# Distinct genetic profiles influence body mass index between infancy and adolescence

**DOI:** 10.1101/2024.07.14.24310392

**Authors:** Geng Wang, Samuel McEwan, Jian Zeng, Mekonnen Haile-Mariam, Loic Yengo, Michael E Goddard, Kathryn E Kemper, Nicole M Warrington

## Abstract

Body mass index (BMI) changes throughout life with age-varying genetic contributions. We aimed to investigate the genetic contribution to BMI across early life using repeated measures from the Avon Longitudinal Study of Parents and Children (ALSPAC) cohort. Random regression modelling was used to estimate the genetic covariance matrix (**K**_**g**_) of BMI trajectories from ages one to 18 years with 65,930 repeated BMI measurements from 6,291 genotyped ALSPAC participants. The **K**_**g**_ matrix was used to estimate SNP-based heritability 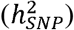 at yearly intervals from 1-18 years and genetic correlations across early life. We also performed an eigenvalue decomposition of **K**_**g**_ to identify age-varying genetic patterns of BMI. Finally, we investigated the impact of a polygenic score derived from adult BMI on the estimated genetic components across early life. The 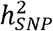 was relatively constant across early life, between 23-30%. The genetic contribution to BMI in early childhood is different to that in later childhood, indicated by the diminishing strength of genetic correlation across different ages. The eigenvalue decomposition revealed that the primary axis of variation (explaining 89% of the genetic variance in **K**_**g**_) increases with age from zero and reaches a plateau in adolescence, while the second eigenfunction (explaining around 9% of **K**_**g**_) represents factors with opposing effects on BMI between early and later ages. Adjusting for the adult BMI polygenic score attenuated the 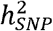 from late childhood; for example, 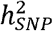 is 29.8% (SE=6.5%) at 18 years of age and attenuates to 14.5% (SE=6.3%) after adjusting for the adult BMI polygenic score. Although common genetic variation explains around 23-30% of BMI variability across early life, our findings indicate that there is a different genetic profile operating during infancy compared to later childhood and adolescence.

## Introduction

Body mass index (BMI), defined as body weight (in kilograms) divided by the square of height (in meters), is a commonly used measure to estimate total body fat. Obesity, defined in adults by BMI exceeding 30kg/m^2^, poses a significant risk for the development of many diseases, particularly cardio-metabolic diseases ^1^. One of the strongest predictors of obesity in adulthood is high BMI during childhood ^2^. In the general population, BMI across childhood involves three distinct phases ^3^. First, there’s a rapid increase in BMI from birth to around nine months, when children reach their adiposity peak (AP). Second, there is a rapid decline in BMI until around five or six years of age where children reach their adiposity rebound (AR), which is due to factors such as changes in body composition and increased physical activity. Finally, BMI gradually increases until early adulthood, reflecting ongoing body development through puberty. Increasing our understanding of the mechanisms influencing BMI across early life could lead to strategies to enable early prevention of obesity.

Considerable strides have been made in understanding the genetic underpinnings of BMI throughout childhood and in adults. The largest and most powerful studies have been conducted in adults including 700K individuals and identifying nearly a thousand independent loci through genome-wide association studies (GWAS) ^4^. The total amount of variation in adult BMI explained by all common genetic variants either on or tagged by GWAS arrays, also known as single nucleotide polymorphism (SNP) based heritability 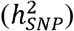, was estimated by Yengo *et al*. (2018) as 22.4% (SE = 3.7%) ^4^. This contrasts to family-based designs which estimate the heritability of BMI in adults to be about 44% (SE = 4%) ^5^, and twin-based estimates which are around 60-80% ^6^. The differences between the experimental designs relate to genetic variation not tagged by common SNP (e.g. rare variants), and potential overestimation of twin-based estimates through common environmental or other effects ^5^. Some studies have also suggested a genotype-by-age interaction for adult BMI for older age groups, implying that the genetic correlation (*r*_*g*_) for BMI between different ages is less than 1 ^5,7^.

Similar to studies in adults, the estimates of heritability of BMI in childhood differ not only on study design but also age. For example, twin studies estimate a heritability of around 40% at age four, presumably where there is a large shared environmental component to BMI ^8^. Estimated heritability from twin-studies increases to around 80% from the age of ten to 19 years old ^8-10^, which is similar to twin-based estimates of heritability in adults (60-80%) ^6^. The estimated SNP-based heritability is 20-40% between one and ten years of age ^9,11,12^, again consistent with the estimate in adults of 22%^4^. BMI in childhood also exhibits genotype-by-age interactions, often to a greater magnitude than is seen in adults. Silventoinen *et al*. pool data from 25 twin cohorts to show that the genetic correlation between BMI at age 4 and 18 years is 0.5 (95% CI 0.38 – 0.61) in males and 0.38 (95% CI 0.24 – 0.50) in females ^13^. Low genetic correlations are also reported using SNP-based estimation where Helgeland *et al*. ^12^ estimated the genetic correlation between BMI at 5 years and in adults to be 0.45 (SE = 0.041) and with Couto Alves *et al*. who estimate the genetic correlation between BMI at the adiposity rebound and adult BMI to be 0.64 (SE = 0.08) ^14^. This means that although BMI is moderately to highly heritable in children and adults, the genetic factors contributing to the heritability are not consistent throughout time and the estimates of heritability in twin-based studies are prone to environmental or other confounders. SNP-based heritability estimates capture only about half the total additive genetic variance but are not as susceptible to confounders.

Helgeland *et al*. identify 46 genomic loci that are associated with BMI in at least one of twelve time points from birth to 8 years of age. Around half of these loci influence BMI during infancy but have no effect after the AR, including no effect on adult BMI. One locus following this pattern of association is *LEP/LEPR*, which has been associated with BMI across early life and at the adiposity peak in numerous studies ^11,12,14^. Additionally, the genetic correlation between the BMI at the AP and adult BMI is lower than the correlation later in childhood (for example, *r*_*g*_= 0.26, SE = 0.07 between BMI at AP and adult BMI^14^ and *r*_*g*_ = 0.63, SE = 0.06 between BMI at 8 years and adult BMI^12^). The age-varying genetic effects at individual loci, along with the genetic correlations estimated to be less than one, indicate that there might be a unique genetic profile influencing BMI during the first few years of life that differs from the genetic profile affecting BMI in adulthood. However, all of these studies use cross-sectional data, which overlook individual-level changes and neglect to utilize the repeated measures data from longitudinal cohorts to explore the evolving genetic profile of BMI trajectories throughout early life.

One method for investigating genetic effects on traits over time using repeated measures data is the random regression model (RRM) ^15,16^. The RRM offers flexibility in modelling the correlation structure of repeated measurements through the inclusion of random effects for each subject and accommodates missing or incomplete data. The method models population, genetic and individual effects as a continuous function of time (e.g. age), thus estimating the population average trajectory as well as random coefficients to describe each individual’s trajectory. Random effects can be partitioned into additive genetic and individual-specific effects, where the additive genetic effect can be estimated using a variance-covariance structure defined by a genetic relationship matrix constructed with common SNPs ^17^. In contrast to previous studies using cross-sectional data, the random regression model increases power to identify patterns of genetic variation by considering the effect of all SNPs simultaneously and it does not attempt to identify individual SNPs associated with a particular time-point or feature of the trajectory. The RRM also reduces the number of parameters estimated from the data compared to previous twin studies which use a series of pairwise comparisons across ages to estimate genetic correlations ^13^. Finally, it is flexible in that genetic correlations can be estimated at any given pair of ages (within the age range of the data).

The aim of the current study is to use repeated measures data from a large birth cohort, the Avon Longitudinal Study of Parents and Children (ALSPAC) cohort, to investigate the genetic profile of BMI across early life and provide insights into whether it differs from BMI in adulthood. We estimate the SNP-based heritability and genetic correlations within the age range of one to 18 years, identify patterns of genetic variation in BMI and explore the influence of an adult BMI polygenic score on the genetic variance of childhood BMI.

## Subjects and methods

### Study Sample

ALSPAC is a population-based, prospective birth cohort conducted in region of Avon in the United Kingdom ^18,19^. Pregnant women resident in Avon with expected dates of delivery between 1st April 1991 and 31st December 1992 were invited to take part in the study. The initial number of pregnancies enrolled was 14,541, with 13,988 children who were alive at 1 year of age. When the oldest children were approximately seven years of age, an attempt was made to bolster the initial sample with eligible families, resulting in an additional 913 children joining the ALSPAC cohort. The total sample size for analyses using any data collected after the age of seven is therefore 15,447 pregnancies, resulting in 15,658 fetuses. Of these 14,901 children were alive at 1 year of age. ALSPAC collected extensive health-related information from the mothers, fathers and their children at regular intervals from birth to early adulthood and blood samples from 9,115 children were collected at various follow-ups for genotyping. Using information supplied by ALSPAC, we restricted our analysis to singleton births and individuals of European ancestry (see details below) ^18,19^. The study website http://www.bristol.ac.uk/alspac/researchers/our-data/) contains details of all the data that is available through a fully searchable data dictionary and variable search tool ^20^.

Ethical approval for the study was obtained from the ALSPAC Ethics and Law Committee and the Local Research Ethics Committees. Informed consent for the use of data collected via questionnaires and clinics was obtained from participants or their mothers following the recommendations of the ALSPAC Ethics and Law Committee at the time. Data access of the ALSPAC cohort can be applied for by submitting a request to the study’s Data Access Committee. Requirements for data access are described at http://www.bristol.ac.uk/alspac/. This project received ethical approval from the Institutional Human Research Ethics Committee, University of Queensland (Approval Number 2019002705).

### Genetic Data

Genotyping, quality control (QC) and imputation procedures were performed centrally by ALSPAC and described in detail elsewhere ^18,19^. In brief, the children were genotyped using the Illumina HumanHap550 quad genome-wide microarray and imputed to the 1000 Genomes reference panel (Version 1, Phase 3, Dec 2013 Release, using haplotypes from all populations) ^21^ using IMPUTE2 (v2.2.2) ^22,23^. Quality control measures applied centrally by ALSPAC included checks for gender mismatches, heterozygosity levels, missingness rates, and Hardy-Weinberg equilibrium. Families who withdrew from the study were removed.

Population stratification was evaluated using multidimensional scaling analysis, which was then compared to HapMap II (release 22) reference populations ^24^, including individuals of European descent (CEU), Han Chinese, Japanese, and Yoruba. The study removed all participants with non-European ancestry centrally. After QC and retaining only individuals of European ancestry from singleton births, there were 8,635 genotyped children with 26,048,419 SNPs. More details of centrally-performed QC are provided at the following webpage: https://proposals.epi.bristol.ac.uk/alspac_omics_data_catalogue.html#.

We applied further QC steps to remove SNPs that displayed more than 5% missingness, a Hardy-Weinberg equilibrium P-value of less than 10^−6^, imputation quality INFO score less than 0.8, and minor allele frequency of less than 1%. A total of 6,380,782 SNPs were retained. Using the cleaned genotype data, we generated a genomic relationship matrix (GRM) using GCTA (version 1.94.1) ^17^. Individuals with a relatedness coefficient of greater than 0.05 were identified and one member of every related pair randomly removed using the ‘--grm-cutoff 0.05’ option in GCTA. This resulted in a set of 7,791 unrelated individuals of European ancestry for further analyses.

### BMI Measurements

The data collected in ALSPAC included height and weight at up to 32 follow-ups before children reached adulthood, including parent and child completed questionnaires, nurse reports from routine health care visits and clinic attendance.

We used the growthcleanr package^25^ in R^26^ to compare each height and weight measurement with a weighted moving average of the individual’s other measurements to identify biologically implausible values in height and weight. Further details about the package are described elsewhere (https://carriedaymont.github.io/growthcleanr/articles/output.html) ^25^. This flagged 5,480 measurements (weight or height [2.1% of the total number of measurements]), from 3,667 individuals, as outliers and we removed them from subsequent analyses. Then, we only retained individuals of European ancestry with genotype information (see ***Genetic Data***). We only used data collected between 1 and 18 years. This avoids the adiposity peak of around 9 months in most individuals meaning that individual trajectories should approximately follow a 3^rd^ order (cubic) polynomial^27^. We excluded measurements after 18 years because the cubic polynomial model indicates a downward curve during early adulthood, where it is expected to remain stable or slightly increase. Finally, we only included individuals with at least four measurements, ensuring there is adequate data per individual to fit the cubic polynomial. The final dataset consisted of 65,930 BMI measurements from 6,291 unrelated individuals (Table 1, an average of 10.5 (SD=3.8) measures per individual). All BMI phenotypes were natural log-transformed for analyses due to skewed distribution of BMI. All data cleaning procedures were performed in R (version 4.2.1).

**Table 1.** Descriptive statistics of the final dataset in ALSPAC cohort used for RRM analysis.

| Categories | ALSPAC follow-up name | N<br>(Male/Female) | Mean age (SD)<br>[years] | Median of BMI (IQR)<br>[kg/m<sup>2</sup>] |
| --- | --- | --- | --- | --- |
| Nurse reports | Child health database 2 | 40 (13/27) | 1.08 (0.07) | 17.14 (2.26) |
|  | Child health database 3 | 4454<br>(2219/2235) | 1.7 (0.23) | 16.8 (1.91) |
|  | Child health database 4 | 4255<br>(2132/2123) | 3.71 (0.2) | 16.15 (1.75) |
| Child in Focus<br>(CIF) * | CIF 12 months | 730 (368/362) | 1.03 (0.02) | 17.84 (1.83) |
|  | CIF 18 months | 721 (372/349) | 1.53 (0.03) | 17.84 (1.83) |
|  | CIF 25 months | 683 (358/325) | 2.08 (0.02) | 17.09 (1.75) |
|  | CIF 31 months | 728 (376/352) | 2.59 (0.02) | 16.74 (1.73) |
|  | CIF 37 months | 723 (374/349) | 3.08 (0.02) | 16.61 (1.61) |
|  | CIF 43 months | 706 (367/339) | 3.59 (0.02) | 16.45 (1.55) |
|  | CIF 49 months | 706 (369/337) | 4.07 (0.03) | 16.36 (1.59) |
|  | CIF 61 months | 694 (361/333) | 5.16 (0.07) | 16.15 (1.55) |
| Clinic<br>measurements | Focus@7 | 5350<br>(2710/2640) | 7.56 (0.29) | 15.81 (2.15) |
|  | Focus@8 | 4651<br>(2332/2319) | 8.68 (0.29) | 16.62 (2.64) |
|  | Focus@9 | 5181<br>(2566/2615) | 9.91 (0.31) | 17.02 (3.35) |
|  | Focus@10 | 5228<br>(2581/2647) | 10.68 (0.24) | 17.5 (3.73) |
|  | Focus@11 | 5072<br>(2498/2574) | 11.78 (0.23) | 18.26 (4.24) |
|  | Teen Focus1 | 4735<br>(2316/2419) | 12.84 (0.22) | 19.05 (4.17) |
|  | Teen Focus2 | 4404<br>(2151/2253) | 13.88 (0.2) | 19.63 (4.03) |
|  | Teen Focus3 | 3885<br>(1867/2018) | 15.48 (0.3) | 20.71 (3.88) |
|  | Teen Focus4 | 3052<br>(1376/1676) | 17.44 (0.25) | 21.81 (4.37) |
| Questionaries | My School Boy/Girl<br>(KN) | 2829<br>(1458/1371) | 5.81 (0.09) | 15.5 (2.06) |
|  | My Daughter/Son<br>Growing up (KP) round 1 | 803 (396/407) | 6.37 (0.35) | 15.63 (2.02) |
|  | My Daughter/Son<br>Growing up (KP) round 2 | 123 (64/59) | 6.34 (0.41) | 15.72 (1.87) |
|  | Your Son/Daughter at 9<br>(KU) round 1 | 783 (397/386) | 9.31 (0.75) | 16.69 (3.23) |
|  | Your Son/Daughter at 9<br>(KU) round 2 | 151 (65/86) | 8.81 (0.7) | 16.4 (2.18) |
|  | Your Son/Daughter at 9<br>(KU) round 3 | 102 (50/52) | 9.08 (0.71) | 16.62 (3.85) |
|  | Your Son/Daughter at 9<br>(KU) round 4 | 59 (27/32) | 9.46 (0.54) | 16.7 (3.35) |
|  | My Teenage<br>Son/Daughter (TA) | 2706<br>(1343/1363) | 13.18 (0.16) | 19.07 (3.89) |
|  | Your Son/Daughter at<br>16+ (TC) | 2375<br>(1220/1155) | 16.87 (0.36) | 20.9 (3.35) |

\* Ten of the clinic follow-ups before 7 years of age were designed to focus on a 10% subset of individuals (termed the “Child in Focus”). N: sample size, SD: standard deviation, IQR: Interquartile range

### Statistical Model

Our random regression models the change in BMI with age as a polynomial in age in which the curve for each person is the sum of an overall age effect, common to everybody, and an individual departure from this curve described by a random genetic effect and a random effect specific to that person. We used Legendre polynomials of standardized age in the RMM ^28^. Legendre polynomials are a system of orthogonal polynomials over the interval [-1, 1] that reduce the correlation among the random regression coefficients, making them computationally efficient for longitudinal modelling. We therefore standardized age (*m*) using the following equation:

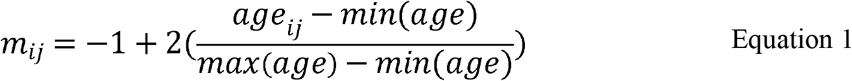

where *age*_*ij*_ is the age of individual *i* at the measurement *j, min*(*age*) is the youngest age (i.e 1 year) and *max*(*age*) is the oldest age (i.e. 18 years) in the data. This rescales the age range from -1 to 1 (rather than 1 to 18) to facilitate the use of the Legendre polynomials.

We used the standardized age to generate a matrix of Legendre polynomials evaluated at specified ages, **Φ**:

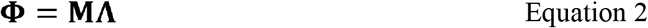

where **Φ**is a matrix of dimension *t* × *k*, where *t* is the standardized age and *k* is the degree of the polynomial plus one (in our case, *k* = 4; for example, coefficients for the overall mean [intercept], linear, quadratic and cubic polynomials of standardized age). ***M*** is a matrix of order t × k containing the standardized age values in their m^k^ form (i.e. m^0^, m^1^, m^2^, m^3^). ***Λ*** is a matrix of Legendre polynomial coefficients of order *k* × *k*, taking the form ^29^:

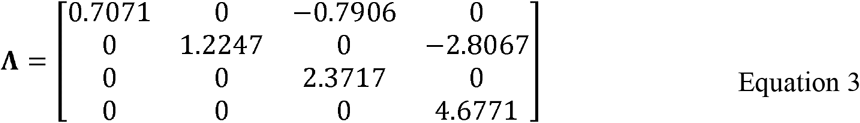

Note we used **Φ**in two different contexts below. First, rows from **Φ**were used in the design matrices of the RRM model when **M** contains the observed ages where BMI was collected. Second, we used regularly spaced ages along the interval [-1, 1] for **M** when transforming variance components from the RRM back onto the observed age-scale.

The following RRM was fitted to the observed data:

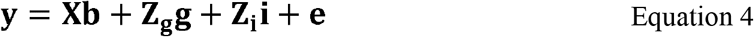

where **y** is a *n* × 1 column vector of log-transformed BMI observations (*n* is the total number of observations). ***X*** is a *n* × (*k*_*f*_ + c) design matrix for the fixed effects, where *k*_*f*_ is the degree of the polynomial plus one in the fixed effects, and c is the number of covariates. The fixed effects include overall mean (or intercept), linear, quadratic and cubic Legendre polynomials, and covariates: sex, interaction terms between sex and the Legendre polynomials, and measurement source (whether the BMI measure was collected via questionnaire or at the clinic [including both the nurse report and ALSPAC clinic visits]). **b** is a (*k*_*f*_ + c) × 1 vector of estimated fixed regression coefficients. **Z**_**g**_ and **Z**_**i**_ are *n* × (*k*_*g*_ × *N*) and *n* × (*k*_*i*_ × *N*) design matrices, respectively, for the random effects, where N is the number of individuals and *k*_*g*_ and *k*_*i*_ is the degree of the polynomial plus one in the random effects (*k*_*g*_ and *k*_*i*_ are less than or equal to *k*_*f*_). **Z**_**g**_ and **Z**_**i**_ contain rows of **Φ**, for the relevant Legendre polynomials at the observed age for an observation. We have specified two unique design matrices, **Z**_**g**_ and **Z**_**i**_, to allow the degree of the polynomial (and therefore the value of *k*_*g*_ and *k*_*i*_) to differ between the additive genetic and unique individual effects. **g** is a (*k*_*g*_ × *N*) × 1 vector of random additive genetic polynomial effects, and **i** is a (*k*_*i*_ × *N*) × 1 vector of random individual-specific effects (i.e., individual-specific effects on BMI not captured by SNP genotypes such as, maternal effects, environmental effects, and genetic factors that are not tagged by common SNPs). Finally, e is a *n* × 1 vector of residuals, with the assumption that the variance of the residual term remains constant over time.

The distribution of random effects, **g, i** and **e**, follow a normal distribution with mean zero and variance given by:

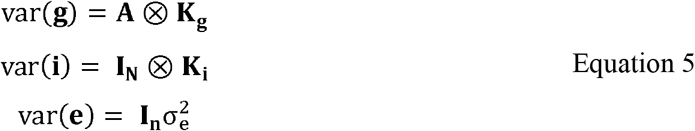

where **A** is the genomic relationship matrix constructed with SNP genotypes, as described above, ⊗ denotes the Kronecker product, **K**_**g**_ and **K**_**i**_ are the *k*_*g*_ × *k*_*g*_ and *k*_*i*_ × *k*_*i*_ variance-covariance matrices for the additive genetic and unique individual effects, respectively; and **I**_**N**_ and **I**_**n**_ are identity matrices with appropriate order for the respective random effects, 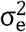 is the variance of residuals. The model was fit using ASReml-SA version 4.2.1 (https://vsni.co.uk/software/asreml), with the ASReml model specification file (.as file) provided in **Supplementary Note 1**.

### Model fitting

We began by fitting cubic Legendre polynomials as fixed effects, and for both the additive genetic and unique individual effects. We examined model fit visually in a subset of randomly selected individuals (**Supplementary Figures 1 and 2**) and inspected the estimated variance components with standard errors. We used the Akaike Information Criterion (AIC) and a likelihood-ratio test (LRT) to formally determine the appropriate degree of polynomial function for the random effects, ensuring an accurate fit to the data (**Supplementary Table 1**).

### Transformation of variance components from the polynomials to the observed scale of one to 18 years

The genetic variance estimated by the RRM, **K**_**g**_, is a *k*_*g*_ × *k*_*g*_ variance-covariance matrix where the elements relate to the genetic variance of the intercept (or mean), slope and quadratic terms (when *k*_*g*_ = 3). To aid in interpretation, these polynomial terms can be transformed back onto the original scale of age using **Φ**, a matrix of Legendre polynomial coefficients (equation 2). As noted above, we can use any arbitrary number of values along the interval [-1, 1] for **M** when calculating **Φ** during this transformation as the variance components from the RRM are defined on a continuous scale. Thus, the estimates of the additive genetic 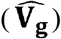 and unique individual 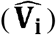 variance-covariance matrices on the observed scale are:

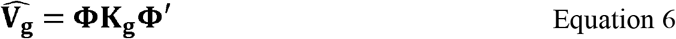

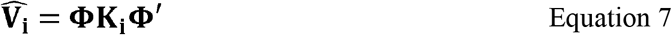

where 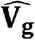 and 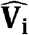 are of order *t* × *t*, and *t* is the number of ages across the BMI trajectory of interest for evaluation. For simplicity, we chose 18 equally spaced values along [-1,1] for **M** corresponding to ages of 1 to 18 years, and thus *t* = 18. The estimates for the phenotypic variance 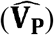, SNP-based heritability 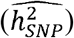, and genetic and phenotypic correlation between time points t_1_ and 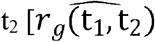 and 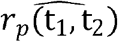, respectively] were calculated as:

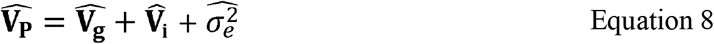

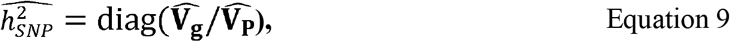

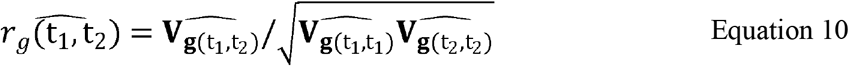

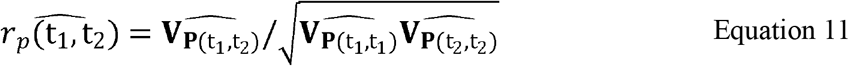

where 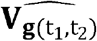 is the (t_1_, t_2_) element from the estimated additive genetic variance-covariance matrix 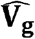 and 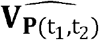 is the (t_1_, t_2_) element from the estimated phenotypic variance-covariance matrix 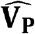. Standard errors for the variance components, SNP-based heritability and genetic correlations were calculated using a Taylor series expansion following Fischer *et al*. ^30^.

### Patterns of genetic variation

We use eigenvalue decomposition on **K**_**g**_ to examine the patterns of genetic variation over the age space ^28,31^. These patterns could be observed in 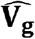 but the eigenvalue decomposition on **K**_**g**_ has the advantage of being independent of the choice of the ages to evaluate in **Φ**. Then,

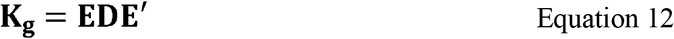

where the matrix **K**_**g**_ (Equation 5) is partitioned into its eigenvalues **D** and eigenvectors **E**, using eigen() function in R. The eigenvectors are transformed into eigenfunctions (of age) as **Λ**E, where **Λ** is a matrix of Legendre polynomials (given in Equation 3), and each column of **Λ**E represents the coefficients for each eigenfunction (*ψ*). Eigenfunctions can also be evaluated at a specific set of ages as **Φ**E.

Since each eigenvector has an associated eigenvalue, we can test whether each eigenfunction explains more than zero genetic variance by using their eigenvalues ^28^. The coefficient matrix (**E**) is restricted by setting *q* testing eigenvalues (e.g. eigenvalue 3) in **D** to zero, resulting in **D**^*^ and 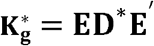. The approximation can then be tested using a χ^2^ test with *q*(*q*+1)/2 degrees of freedom as:

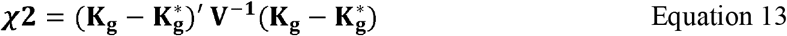

where **V**^-**1**^ is the inverse of the sampling variance-covariance matrix for **K**_**g**_ from the fitted model (and can be obtained from the .vvp file from ASReml). We constructed 95% confidence intervals for the eigenvalues using numerical simulation (see **Supplementary Note 2**).

### Evaluating BMI trajectories using the eigenfunctions

To further assess the eigenfunctions, we calculated a polygenic score (PGS) for each eigenfunction and evaluated the BMI trajectory for low (<1 SD from the mean), average (within 1 SD of mean) and high (>1 SD from the mean) eigenfunction PGS. In this context, the PGS evaluates the sum of additive genetic effects tagged by common SNP for the primary or secondary axes of genetic variation (i.e. eigenfunction). The PGS for eigenfunctions on the original (age) scale as:

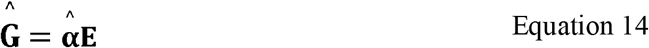

where **Ĝ** is a *N* × *k*_*g*_ matrix of the polygenic scores of the *N* individuals for eigenfunction *i* (*i* = 1 or 2), 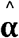 is a *N* × *k*_*g*_ matrix of estimated random regression coefficients for the Legendre polynomials for the additive genetic effects and **E** is the matrix of eigenvectors (Equation 12).

All analyses and plotting were performed in R (version 4.2.1). P values of fixed effects and random variances were calculated using the the chi-squared distribution function with one degree of freedom (**Supplementary Table 2 and 3**).

### Adjusting for adult BMI PGS

Next, we explored whether the SNP-based heritability across childhood was explained by the genetic variants known to be associated with adult BMI. We downloaded the SNP effect size estimates on adult BMI for approximately 7 million common SNPs generated from a SBayesRC^32^ analysis of GWAS data of adult BMI in unrelated individuals of European ancestry (N= 347,800, age ≥ 40 years) from the UK Biobank^33,34^ (https://sbayes.pctgplots.cloud.edu.au/data/SBayesRC/share/v1.0/PGS/). We constructed the adult BMI PGS in our cohort using the quality-controlled genetic data (see ***Genetic Data*** section) and the --score sum option in Plink v 1.9 ^35^, which creates a sum of SNPs weighted by the adult BMI effect size. Subsequently, we integrated the adult BMI PGS and its interaction with age (i.e. interaction terms between PGS and the linear, quadratic, and cubic Legendre polynomials) as covariates in the fixed effects part of our RRM. The ASReml model specification file (.as file) is provided in **Supplementary Note 3**.

### Secondary analyses

We conducted secondary analyses to ensure our interpretation of the SNP-based heritability, genetic correlations and genetic profiles estimated using the RRM were consistent with other approaches.

First, we estimated SNP-based heritability and genetic correlations in GCTA (version 1.94.1) ^17^ using selected measurements of BMI at cross-sectional follow-ups (i.e. average ages are 0.8 (closest cross-sectional time point to one year available), 1.7, 7.6, 10.7, 13.9, 15.5, and 17.5 years). Each analysis included only one measurement per individual, and included sex and age at measurement as covariates. We mostly used the BMI measurements of the same individuals included in the RRM analyses but included some additional records from the Child health database 2 (mean age =0.8 year, N=4799) and Teen Focus 4 (mean age =17 years, N=3373), which were excluded from the RRM as they were outside 1-18-year age range.

Second, our RRM (Equation 4) assumes that the variance of the residual term is constant over ages. However, the phenotypic variance of BMI increases with age, and therefore the variance of the residual term may also vary with age. This may introduce bias into the estimation of both additive genetic and unique individual variances. In order to address this concern, we conducted a secondary analysis in which we assigned different residual terms for each year in our model allowing them to be estimated (i.e. fitting 17 residual terms in the random regression model). The ASReml model specification file (.as file) is provided in **Supplementary Note 4**.

Third, we performed eigenvalue decomposition using the genetic correlation matrix of BMI obtained from the previously published COADTwins project ^13^, which was described in **Supplementary Note 5**.

## Results

In the RRM analysis, model comparison indicated that the model with a quadratic slope in additive genetic component (*k*_*g*_ = 3) was the superior fit to the data, over fitting a cubic or linear slope (**Supplementary Table 1**). The fixed effects, which are all associated with log(BMI) (P <0.05), are presented in **Supplementary Table 2**, and the estimated covariance matrices for random effects (*K*_*g*_ and *K*_*i*_) and residual variance 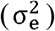 are presented in **Supplementary Table 3**. The examination of estimated trajectories and actual measurements of randomly selected individuals (**Supplementary Figures 1** and **2**) indicates model is a good fit in general. Since the cubic polynomial term for the additive genetic effects were not significant, we used AIC and LRT to formally compare models with (up to) cubic or quadratic degree of the polynomials for the additive genetic effects. The final model retained quadratic polynomials in additive genetic component (*k*_*g*_ = 3), cubic polynomials to model random individual-specific effects (*k*_*i*_ = 4), and cubic polynomials for each sex to model the overall population BMI trajectory.

### Genetic variance and SNP-based heritability

Additive genetic variation was observed for the intercept (**K**_**g 1**,**1**_ =0.0073, SE=0.0013, **Supplementary Table 3**), linear slope (**K**_**g 2**,**2**_=0.0017, SE=0.0003), and quadratic slope (**K**_**g 3**,**3**_ 0.0004, SE=0.0001), indicating that genetics influence on average BMI as well as the shape of BMI trajectory over time. We detected a positive genetic correlation between the intercept and the linear slope (0.682, SE=0.072), and a negative genetic correlation between quadratic slope and both the intercept (−0.678, SE=0.132) and the linear slope (−0.473, SE= 0.174). The estimated SNP-based heritability of the intercept (28.4%, SE=4.8%), linear slope (23.8%, SE=4.2%) and quadratic slope (9.8%, SE=3.1%) were all significantly different from zero (P < 0.05).

We estimated phenotypic, additive genetic and unique individual variance over time between one and 18 years of age on the observed age-scale (**Figure 1**). We observed that variance components tended to increase with age. The SNP-based heritability was significantly greater than zero and ranged between 23-30% across all ages, which is consistent with the SNP-based heritability estimate for the intercept (i.e. average age 9.5 years, **Figure 1 and Supplementary Table 4**).

**Figure 1:**
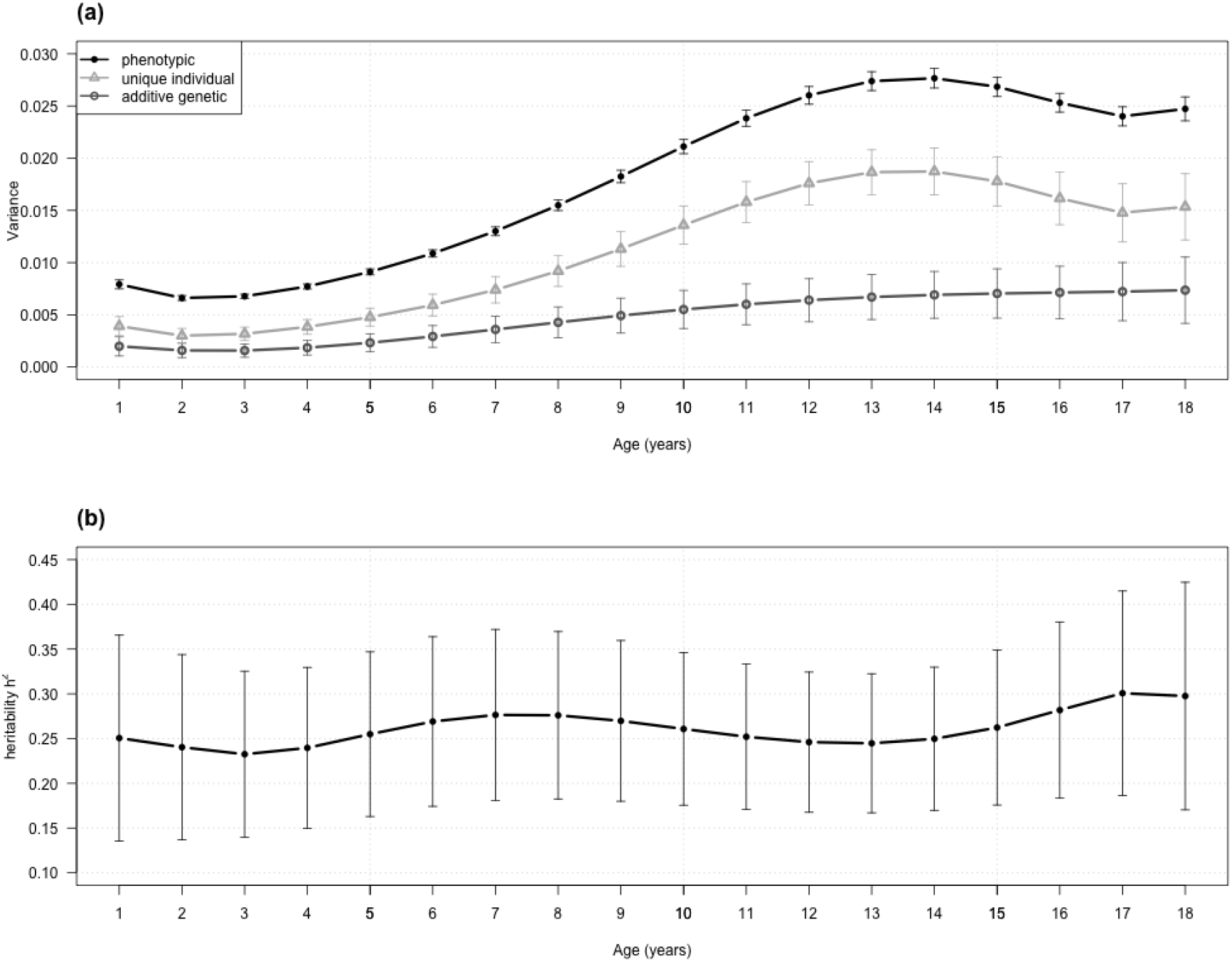
Estimated phenotypic, SNP-based additive genetic and unique individual variance components (a) and SNP-based heritability (b) from one to 18 years of age in the ALSPAC cohort. The error bars indicate 95% confidence intervals.

### Phenotypic and genetic correlations

The genetic correlation (*r*_*g*_) between BMI at two different ages decreases as the difference between the ages increases (**Figure 2, Supplementary Figure 3, Supplementary Table 5**). For example, the genetic correlation between one and two years of age was 0.948 (SE = 0.015), whereas the genetic correlation between one and 10 years was not significantly different from zero (*r*_*g*_ = -0.009, SE = 0.142). Additionally, the genetic contribution to BMI in early childhood (up until approximately 6 years of age) appears to be independent of genetic influences in later childhood and adolescence. This can be seen by the 95% confidence intervals around the estimates of genetic correlation of BMI between age one and from age seven crossing zero. In contrast, the phenotypic correlation of BMI between age one and the subsequent ages decays quicker but remains non-zero (**Supplementary Figure 4, Supplementary Table 6**). For example, the phenotypic correlation between one and two years of age was 0.67 (SE=0.01), whereas the phenotypic correlation between one and 10 years was 0.19 (SE=0.01). Interestingly, the genetic correlations at subsequent ages are higher than the phenotypic correlations, indicated by the relatively smooth curves in the genetic correlation figures (**Figure 2** and **Supplementary Figure 3**) but the sharp peaks in the phenotypic correlation figures (**Supplementary Figure 4**).

**Figure 2:**
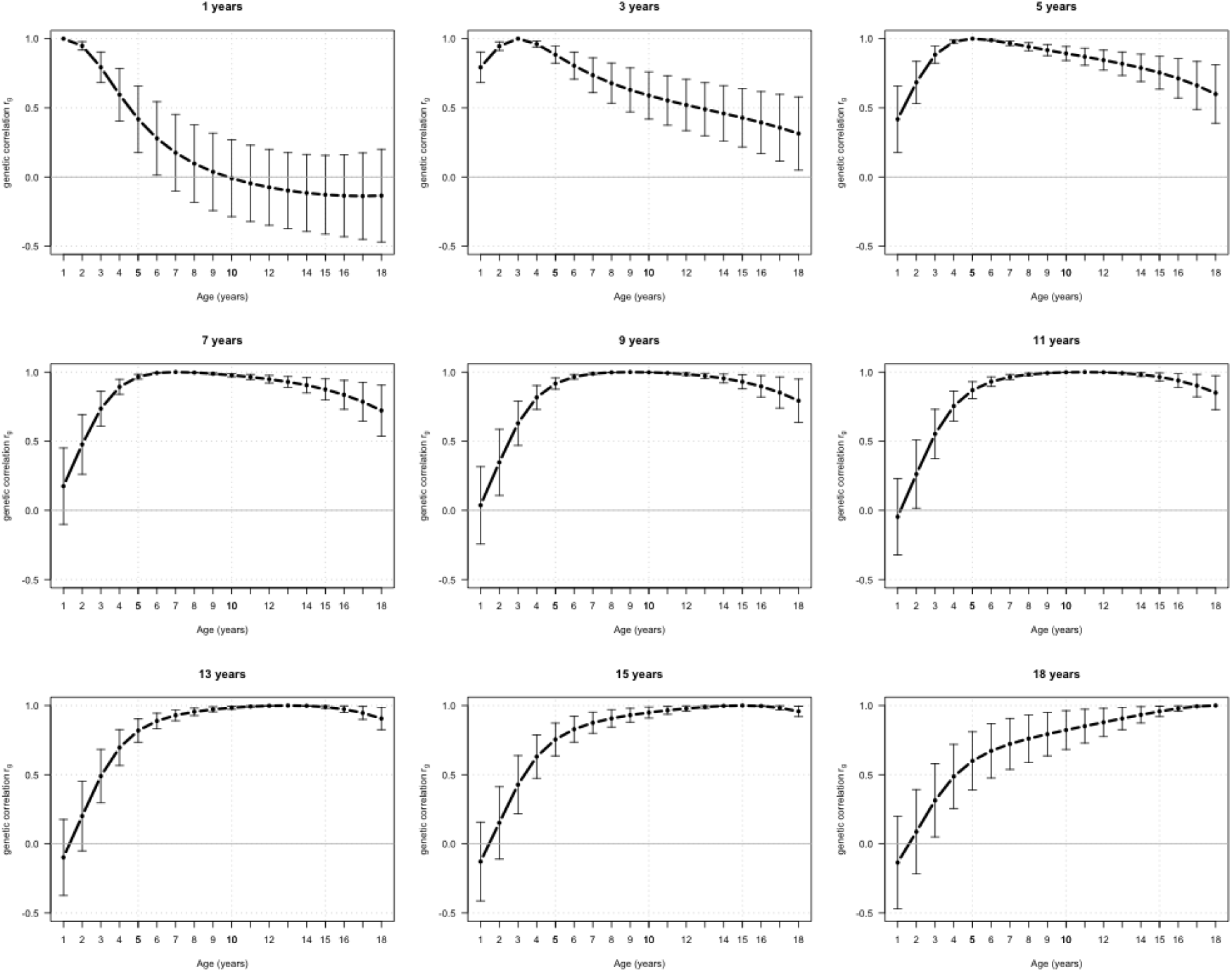
Estimates of genetic correlation (*r*_*g*_) of BMI between different ages from one to 18 years. Each plot represents the genetic correlations (y-axis) between a single age (described above the panel) and all other ages between 1 and 18 years (x-axis). Error bars represent the 95% confidence intervals. The grey horizontal line indicates zero genetic correlation.

### Patterns of genetic variation

We conducted an eigenvalue decomposition on **K**_**g**_ to obtain the eigenvalues 1-3 (i.e.,) and their associated eigenfunctions (,,), where the eigenfunctions represent independent (uncorrelated) axes of genetic variation. The eigenvalues showed that the 1^st^ eigenfunction, *ψ*_1_, accounts for the majority of the variance in **K**_**g**_ (approximately 89%). The 2^nd^ eigenfunction, *ψ*_2_, explains approximately 9% of the variance in **K**_**g**_, while the 3^rd^ third eigenfunction, *ψ*_3_, explains around 2% of the variance in **K**_**g**_. The chi-squared test showed that the 3^rd^ eigenvalue was not significantly different from zero (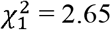, P = 0.10), but that the 2^nd^ eigenfunction did explain a significant proportion of the variance in **K**_**g**_ (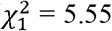, P = 0.02). Numerical simulation of the 95% CI for the eigenvalues supports these findings (**Supplementary Figure 5**). The first and second eigenfunctions were given by:

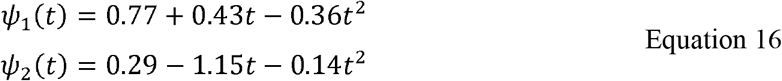

where *t* is the standardized age. From the coefficients above, we can see a relatively large weight of the intercept and quadratic slope in *ψ*_1_ in comparison to the corresponding weights for *ψ*_2_. While *ψ*_2_ is dominated by a relatively large negative weight on the linear slope term.

Evaluation of the eigenfunction across the 1-18 age range shows that *ψ*_1_ monotonically increases from zero over time until it reaches an approximate plateau at around 10 years of age (**Figure 3**). It should be noted that the eigenfunctions indicate an axis of genetic variation, and the key feature of the eigenfunction is its relative change and its relationship to zero. Thus, *ψ*_1_ is always above the x-axis, indicating that it represents positive genetic covariance in BMI across all ages, increasing in strength through childhood and then plateaus at adolescence. In contrast, *ψ*_2_ decreases approximately linearly and crosses zero at approximately 11.5 years. This indicates *ψ*_2_ represents genetic variation with strong positive genetic covariance in infancy, but negative genetic covariance between infancy and later life.

**Figure 3:**
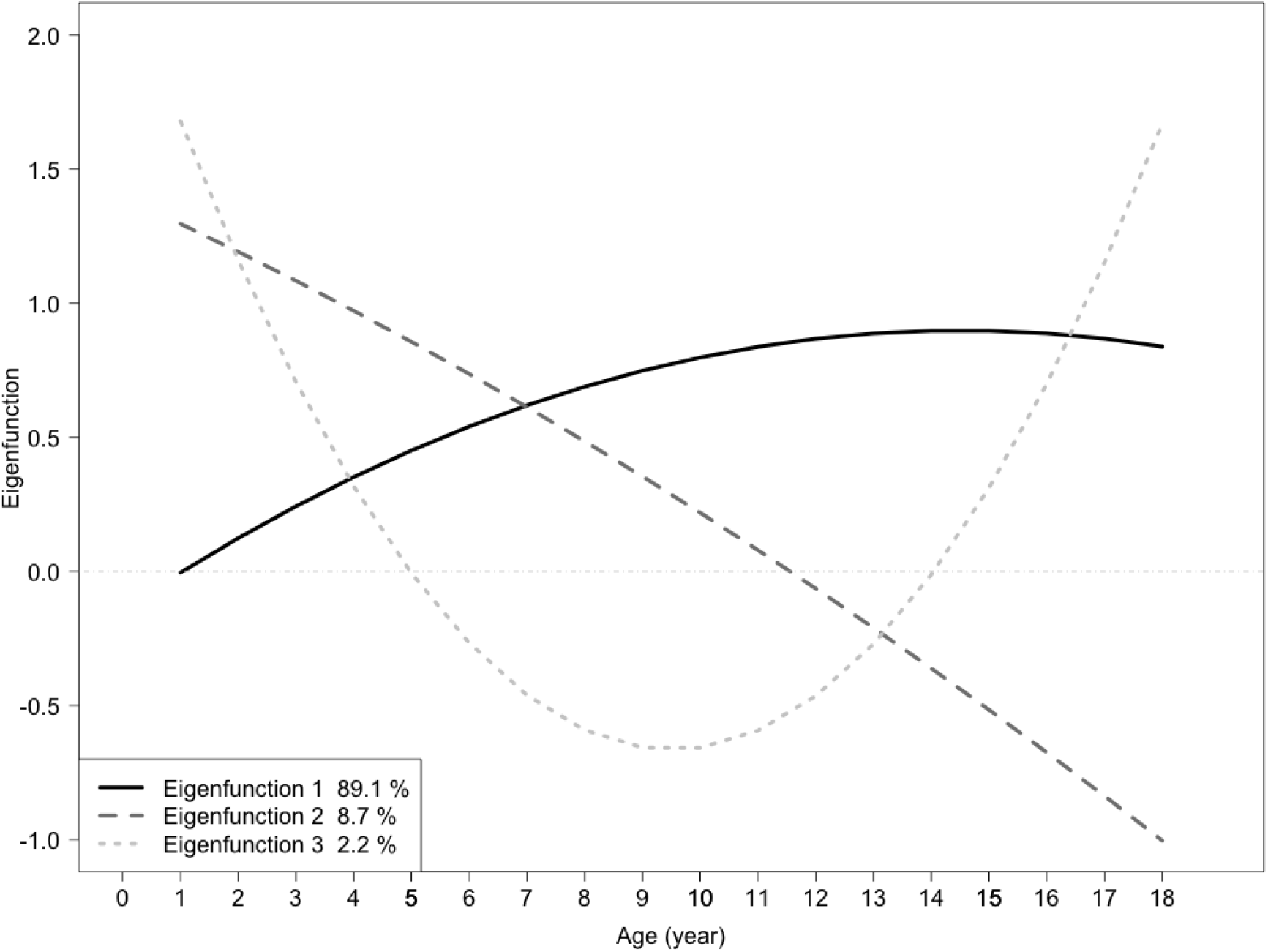
Eigenfunctions of additive genetic effects on BMI evaluated at 1 to 18 years of age. This figure illustrates the eigenfunctions representing the additive genetic effects on BMI as age changes. The x-axis denotes age in years, while the y-axis represents the value of the eigenfunctions. Eigenfunctions 1, 2, and 3 are represented in lines in grayscale (black, dark and light grey, respectively).

To investigate the properties of the eigenfunctions further, we generated PGS for eigenfunctions 1 and 2 and used them to categorise individuals into three clusters: those within one standard deviation of the mean PGS, and the remaining individuals with either high (greater than one standard deviation above the mean) or low (greater than one standard deviation below the mean) PGS scores. **Figure 4** illustrates the mean BMI at each age and the distribution of individual trajectories for these three clusters using the PGS of eigenfunction 1. Individuals with high PGS for eigenfunction 1 have a higher mean BMI, a steeper slope and a lack of adiposity rebound (usually around 6 years in non-obese children) in comparison to those with average PGS. In contrast, individuals with low PGS for eigenfunction 1 seem to follow more closely to a typical trajectory for BMI, but with a lower mean BMI and a slower increase in BMI after adiposity rebound.

**Figure 4:**
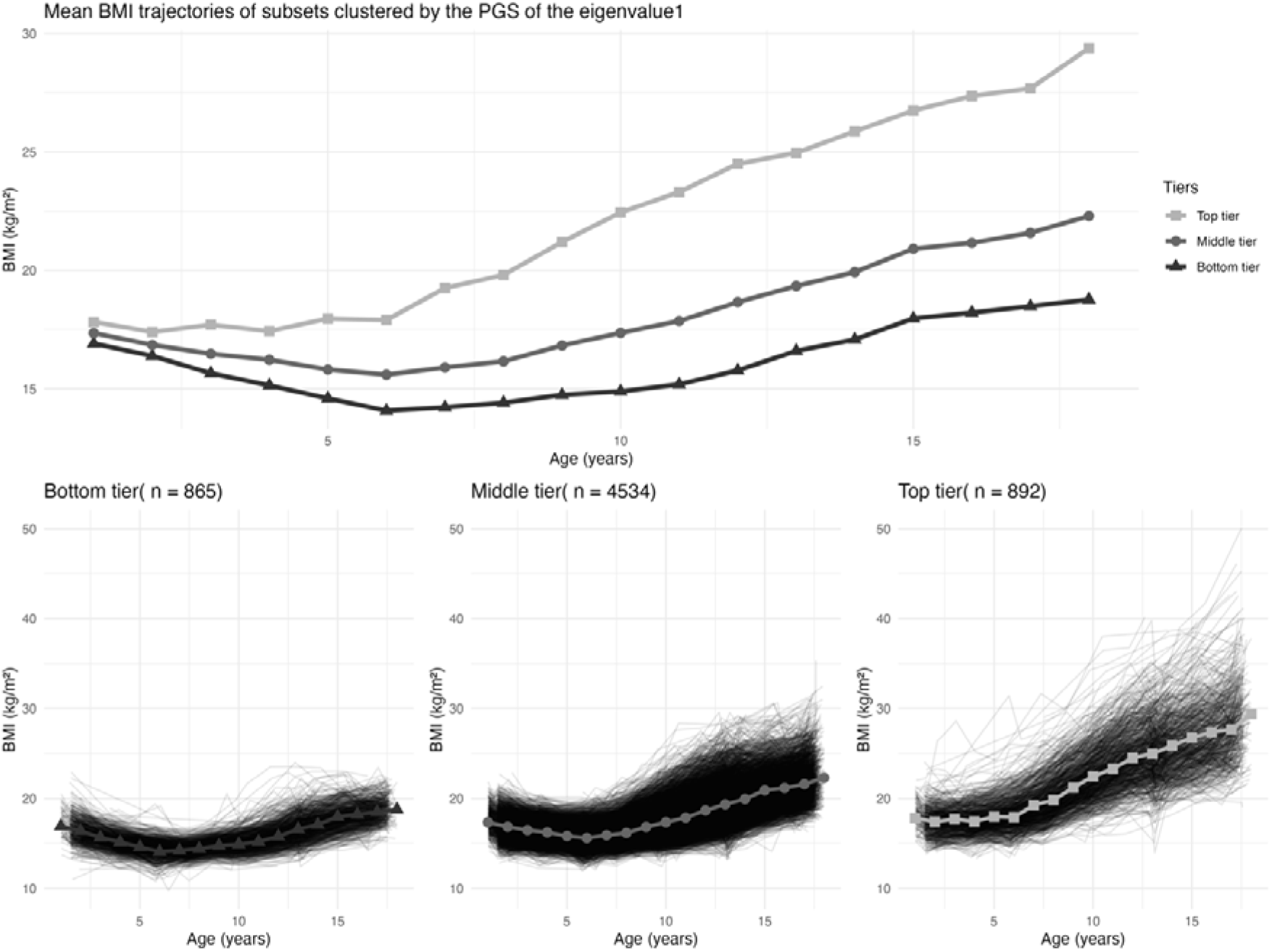
Mean body mass index (BMI) from one to 18 years of age for three clusters based on a polygenic score (PGS) of eigenfunction one. The upper plot illustrates the mean BMI from age one to 18 years for each of the three clusters classified by PGS of eigenfunction one: the top tier are those individuals greater than one standard deviation higher than the mean PGS, the middle tier are those individuals within one standard deviation of the mean PGS, and the bottom tier are those individuals greater than one standard deviation lower than the mean PGS. The lower plots display the same mean BMI as in the upper plot from one to 18 years for the cluster (grey lines) and BMI trajectories for each individual within the three clusters (black lines).

A similar clustering analysis using PGS for eigenfunction 2 (**Supplementary Figure 6**) shows individuals with high PGS have a higher mean BMI at one year, but a flatter trajectory across childhood resulting in a lower mean BMI at 18 years compared to the average PGS and low PGS groups. This reflects the positive genetic effect on BMI represented by eigenfunction 2 in early developmental stages but the opposing effect in later stages.

### Adjustment of childhood BMI for adult BMI polygenic score (PGS)

In the RRM analyses adjusting for an adult BMI PGS, we investigated whether different genetic factors influence BMI in early life compared to mid-to-late adulthood. (**Supplementary Tables 7 and 8, Supplementary Figure 7**). By age 18, the SNP-based heritability roughly halved after adjusting for the adult BMI PGS, reducing from 0.298 (SE=0.065) at 18 in the unadjusted analysis to 0.145 (SE=0.063) **(Figure 5** and **Supplementary Table 9**). The 95% confidence intervals at all ages from one to 18 years differed from zero but decreased with increasing age.

**Figure 5:**
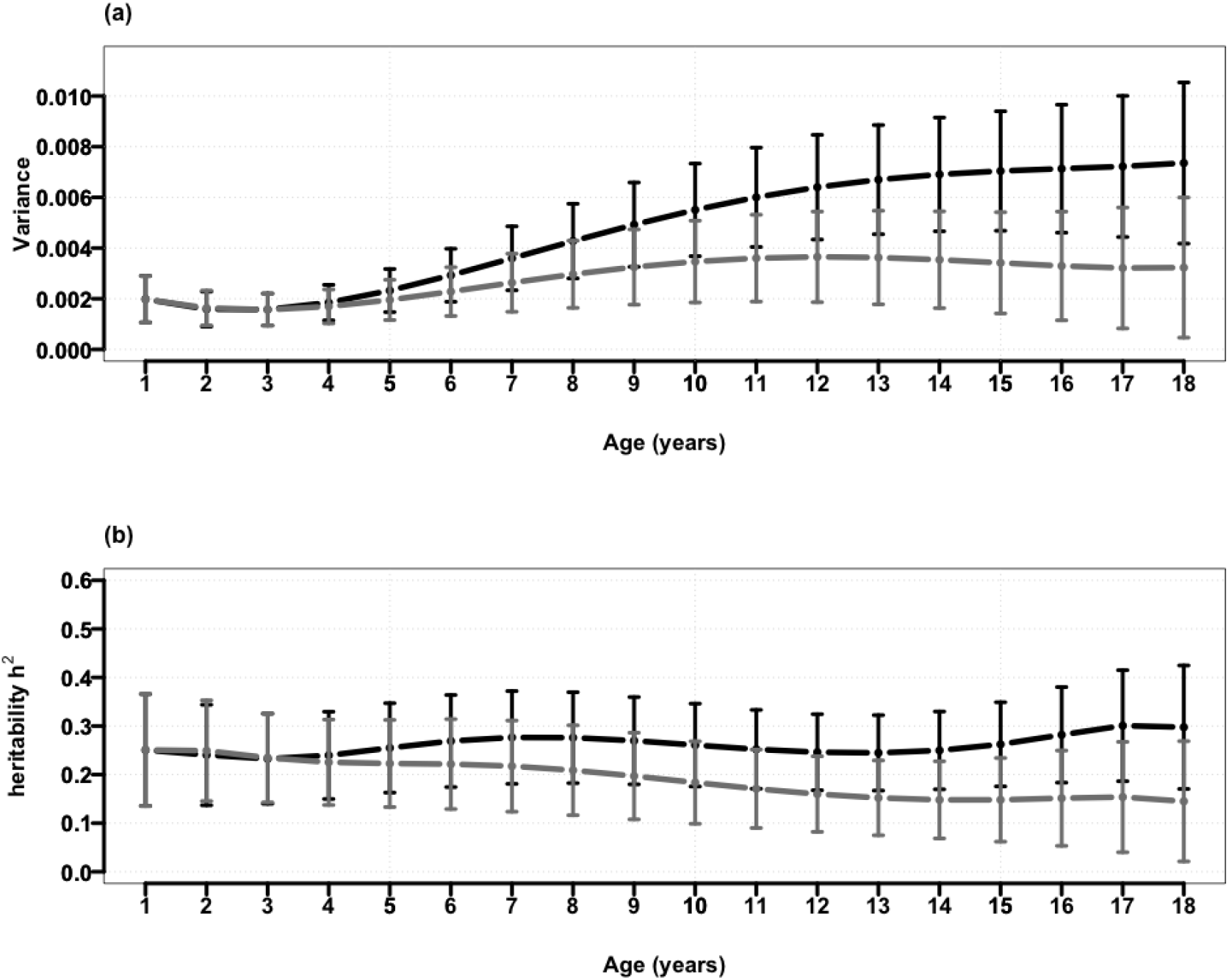
SNP-based heritability and genetic variances with and without adjusting for polygenic score of adult body mass index (BMI). **(a)** additive genetic variance by age, with (grey) and without (black) adjusting for a polygenic score (PGS) of adult BMI in the fixed effects of the RRM; **(b)** heritability estimates of BMI by, with (grey) and without (black) adjusting for a PGS of adult BMI in the fixed effects of the RRM.

### Secondary analyses

#### Cross-sectional analysis

The estimated SNP-based heritability from the RRM were consistent with those from cross-sectional genetic analyses in GCTA, which ranged from 0.28 (SE=0.07) at 0.8 years to 0.37 (SE=0.10) at 17.5 years; however, the standard errors were larger in the cross-sectional analysis than in the RRM model, as expected (**Supplementary Table 10**). Similarly, the genetic correlations estimated using the cross-sectional data in GCTA were similar to the decreasing age-to-age genetic correlations pattern seen in the RRM analyses (**Supplementary Table 11**). For instance, the estimated genetic correlation between Child health database 3 (mean age = 1.7 years) and Focus@7 (mean age = 7.6 years) was estimated at 0.68 (SE=0.15), which was not different to the RRM estimated genetic correlation between 2 and 8 years (rg = 0.4, SE=0.12).

#### Heterogeneous error variance

Estimates of the unique individual and additive genetic variance using the RMM model assuming heterogeneous error variance were similar to those obtained from primary analysis (i.e. assuming homogenous error variance), that is the estimates fall within each other’s 95% confidence intervals (**Supplementary Figure 8, Supplementary Tables 12-14**). This indicates that while there may be some variation in the residuals over time, it does not appear to significantly impact our overall conclusions.

#### Validation of growth patterns

In the eigenvalue decomposition of the genetic covariance matrix of BMI from the COADTwins project, the top eigenfunctions exhibit a similar pattern from one to 18 years old to our findings in ALSPAC in terms of variance explained in BMI (for example, eigenfunction 1 explained ∼89% of the variance in both the ALSPAC and COADTwins) and the shape of the eigenfunctions (eigenfunction 1 was monotonically increasing and eigenfunction 2 decreases over time and crosses zero during adolescence) (**Supplementary Figure 9**).

## Discussion

In the current study, we used a RRM to characterise the genetic profile of BMI from infancy to early adulthood using the ALSPAC cohort. We found significant additive genetic variation affecting the shape of the BMI trajectory from one to 18 years. In other words, there was genetic variation in the mean (or intercept, i.e 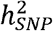 intercept = 28.4%, SE=4.8%) of the BMI trajectory as well as the parameters describing its shape (i.e. 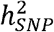 linear slope = 23.8%, SE= 4.2%; and 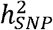 quadratic slope = 9.8%, SE= 3.1%). Since the interpretation of these shape parameters can be tricky, we transformed our estimates back onto the observed age-scale. There was a simultaneous increase in both additive genetic variance and unique individual variance over time, resulting in relatively constant SNP-based heritability 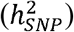 across early life, ranging from 23 – 30%, and not significantly different from the heritability estimated for the intercept. Genetic correlations found that BMI in early childhood is genetically different to BMI in later childhood. Additionally, the SNP-based heritability in early childhood was relatively unaffected by adjusting for an adult BMI PGS, whereas the SNP-based heritability in later childhood attenuated, indicating further that there is a unique genetic profile operating in early life. Finally, the eigenvalue decomposition of the genetic variance-covariance matrix, **K**_**g**_, indicated that the main axis of the genetic variation in BMI throughout childhood is strongly influenced by the mean (or intercept) BMI and progressively amplified over time.

Our study demonstrates genetic variation contributing to childhood BMI trajectories. In clinical settings, pediatricians are primarily concerned with individuals who are underweight (often defined as being less than the 5^th^ percentile on the Centers for Disease Control and Prevention (CDC) percentile charts^36^) or obese (often defined as being greater than the 95^th^ percentile ^36^) The CDC BMI-for-age percentile chart shows that the higher percentiles also have a greater rate of growth from two to 18 years of age. Here, we ascribe some of the variability in growth curve percentiles to genetic factors. We also note a relatively strong genetic correlation between the mean and linear rate of change (i.e. the genetic correlation between the intercept and slope in our RRM is 0.682 [SE=0.072)), which suggests that the correlation between BMI at mean age (i.e. 9.5 years) and rate of change in BMI across childhood can be partly explained by genetics.

Our observed SNP-based heritability estimates 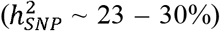 were consistent with the SNP-based heritability for adult BMI ^4^ 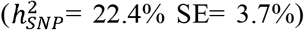 and also the Norwegian Mother, Father and Child Cohort Study (MoBa) in infancy and early childhood ^11,12^ (*h*^*2*^_*SNP*_ ∼ 30 – 40%). In the CODATwins study ^8^, they observed an increase in the heritability from 4 years (42%, 95% CI 37 – 47% in boys; and 41%, 95% CI 35 – 46% in girls) to 19 years of age (75%, 95% CI 67 – 80% in boys; and 75%, 95% CI 67 – 82% in girls). Although we expect the heritability estimates from twin designs to be 3-4 times the SNP-based estimates^5^, our data does not support an increase in heritability throughout this period. These differences could arise from a relative increase in the importance of rare genetic variants influencing BMI, i.e. variants not captured by common SNPs, or other factors associated with twin designs ^5^. We observed that the additive genetic variance captured by common SNPs increased throughout childhood from 0.0020 (SE=0.00047) at year one to 0.0074 (SE= 0.0016) at 18 years. However, there was also a simultaneous increase in unique individual variance and, thus, no significant change in the SNP-based heritability over time.

Our results show decreasing age-to-age genetic correlations as the difference between ages increased from one to 18 years, which is consistent with the CODATwins study ^13^. These results suggested independent genetic influences affecting BMI in infancy and early childhood compared to later childhood and adolescence. Our findings align with the transient genetic effects of SNPs detected in early life but not later life, as found in previous studies (e.g., *LEPR* ^12,14^), which suggests that larger GWAS are needed to finely map the genetic profile of BMI during early stages.

While genetic correlation focuses specifically on shared genetic influences between ages, phenotypic correlation captures overall associations between ages, regardless of their underlying causes. Previous studies investigating phenotypic risks for childhood obesity, such as Geserick *et al*. 2018 ^37^, have highlighted weight gain between two and six years as a key predictor of obesity in adolescence. Our study found that the phenotypic correlation between BMI at 18 years of age and all earlier ages progressively increased from *r*_*p*_ = 0.16 (SE=0.021) at age one to *r*_*p*_ = 0.88 (SE=0.003) at age 17, indicating that high BMI between ages two and six years might not be the only key time point for predicting obesity in adolescence. Interestingly, the phenotypic correlations were weaker than the (SNP-based) genetic correlations among nearby ages (*r*_*p*_=0.67 [SE=0.01], *r*_*g*_=0.95 [SE=0.02] between one to two years) but demonstrated greater strength in ages further apart (*r*_*p*_=0.16 [SE=0.02], *r*_*g*_=-0.14 [SE=0.17] between one to 18 years). The genetic correlations are higher than phenotypic correlations between ages that are close together but lower for ages that are far apart. As the age difference narrows, the model predicts that the genetic correlation approaches 1.0 whereas the phenotypic correlation does not because all measurements are subject to a residual effect and these are all independent. The lower genetic correlation than phenotypic correlation when the ages are far apart implies that genetic effects on BMI are more age specific than environmental effects on BMI.

The eigenvalue decomposition of the genetic variance-covariance matrix allows us to explore the proposed model by Couto Alves *et al*. of genetic influences on childhood BMI^14^. This model, based on GWAS of key BMI trajectory features such as BMI at the adiposity peak and rebound, proposes two distinct biological factors underlying childhood BMI. The first acts primarily in infancy until roughly 8 years of age, and the second acts from birth with increasing strength until 18 years. Our results confirm that the majority of genetic variance in childhood BMI (∼89%) is associated with a factor strongly influenced by the BMI at the mean age (i.e. age 9.5), and which is progressively amplified throughout childhood and adolescence. This factor does not have a strong influence on BMI during infancy. Our results also indicate a second (orthogonal) axis of variation that acts primarily during infancy with decreasing importance during childhood, and a negative covariance during adolescence. Therefore, by using eigenvalue decomposition on our RRM, we were able to provide stronger evidence for the proposed model of Couto Alves *et al*. (2019) by defining two statistically independent axes of variation influencing childhood BMI and determining the degree of genetic variance associated with each axis.

Finally, we performed an analysis adjusting for an adult BMI PGS to (partially) account for genetic factors influencing adult BMI in our analysis. Zheng *et al*. found the PGS explained 16% of the variance in adult BMI ^32^. We observed a similar magnitude of attenuation in the estimated SNP-based heritability at 18 years old (by about 15%) after adjustment of the PGS, which is consistent with the adult PGS being an imperfect predictor of the genetic variance tagged by common SNP in adults. These results were also in line with the genetic correlation and eigenvalue decomposition results, whereby adjustment for the adult BMI PGS did not influence the variance components or SNP-based heritability during infancy (< 3 years). This indicates that the genetic contribution to BMI during childhood to adolescence is shared with that of mid-to-late adulthood and highlights the unique genetic underpinnings of BMI during the infancy period.

The strengths of the current study include the utilisation of ALSPAC, a comprehensive long-term birth cohort that provides valuable insights into BMI trajectories across childhood development. The use of the RRM allowed us to estimate the genetic variance components at any age on the trajectory. It also leveraged the large number of repeated measurements per individual (an average of 8 BMI measures per individual), improving the precision of our estimates over a traditional cross-sectional approach. Additionally, by applying eigenvalue decomposition of the RRM, we revealed patterns of genetic variation in BMI that change over time and for the first time validated the proposed model of childhood BMI by Couto-Alves and colleagues. However, there are several limitations in our study also. Firstly, our estimates of the variance components and SNP-based heritability might be imprecise due to sample size, in comparison to those by the larger MoBa^12^ and CODATwins cohorts^8^, and we were therefore unable to identify fluctuations in SNP-based heritability across childhood. However, RRM estimates have narrower confidence intervals for genetic variation than cross-sectional studies with the same cohort size and can estimate genetic variation at any time point, even without direct data collection. Secondly, the non-significant low or negative genetic correlation in BMI between ages further apart should be interpreted with caution, as genetic correlations are estimated with lower accuracy compared to phenotypic correlations, especially when the age intervals are large. Thirdly, although the overall fitting of the Legendre polynomial function is satisfactory, improvements in model fitting may be possible by exploring other functions for age within the random regression framework (e.g. splines).

In summary, the current study shows that there is a strong genetic drive regulating BMI during childhood and adolescence. Investigating the genetics of BMI during infancy is likely to identify genetic variants that differ from loci associated with adult BMI. Our findings provide justification for exploring the genetics of childhood BMI (e.g. through GWAS), as it is likely to discover genetic variants distinct from those loci associated with adult BMI. It also highlights the presence of age-varying genetic effects on BMI, emphasising the need to consider these factors when studying the genetics of childhood BMI, its relationship to adult BMI and obesity risk.

## Data Availability

The Avon Longitudinal Study of Parents and Children cohort can be applied for by submitting a request to the Data Access Committee. Requirements for data access are described at http://www.bristol.ac.uk/alspac/.

## Appendices

See supplementary materials.

## Declaration of interests

The authors declare no competing interests.

## Acknowledgement

This research has been conducted using the ALSPAC (Reference B4194). We are extremely grateful to all the families who took part in this study, the midwives for their help in recruiting them, and the whole ALSPAC team, which includes interviewers, computer and laboratory technicians, clerical workers, research scientists, volunteers, managers, receptionists and nurses. We would like to express our sincere gratitude to Dominic Waters (The University of New England, Armidale, Australia) for provision of code from Waters *et al*. (2022)^38^ for calculating standard errors for the parameter estimates. This publication is the work of the authors, and N.M.W will serve as the guarantor for the contents of this paper.

## Funding

G.W. and N.M.W. are funded by a National Health and Medical Research Council (Australia) Investigator grant (APP2008723). K.E.K. was supported by the Australian Research Council (grant FL180100072). The UK Medical Research Council and Wellcome (Grant ref: 217065/Z/19/Z) and the University of Bristol provide core support for ALSPAC. GWAS data in ALSPAC was generated by Sample Logistics and Genotyping Facilities at Wellcome Sanger Institute and LabCorp (Laboratory Corporation of America) using support from 23andMe. A comprehensive list of funding provided to ALSPC from grants is available on the ALSPAC website (http://www.bristol.ac.uk/alspac/external/documents/grant-acknowledgements.pdf). This research was specifically funded by Wellcome Trust and MRC (core), 076467/Z/05/Z.

## Data and code availability

The dataset supporting the current study have not been deposited in a public repository because ALSPAC access policy but are available upon application via https://www.bristol.ac.uk/alspac/researchers/access/. The all code generated or analysed during this study are included in the published article or available from the corresponding author on request.

## Notes

### Competing Interest Statement

The authors have declared no competing interest.

### Author Declarations

The Avon Longitudinal Study of Parents and Children Ethics and Law Committee and the Local Research Ethics Committees gave ethical approval for this work. The Institutional Human Research Ethics Committee of the University of Queensland gave ethical approval for this work.

## References

1. World Health Organization (2021). World Health Statistics 2021: Monitoring health for the SDGs, sustainable development goals. https://www.who.int/data/gho/publications/world-health-statistics.

2. Woo, J.G., Zhang, N., Fenchel, M., Jacobs, D.R., Jr., Hu, T., Urbina, E.M., Burns, T.L., Raitakari, O., Steinberger, J., Bazzano, L., et al. (2020). Prediction of adult class II/III obesity from childhood BMI: the i3C consortium. Int J Obes (Lond) 44, 1164–1172. 10.1038/s41366-019-0461-6.

3. Rolland-Cachera, M.F., Deheeger, M., Bellisle, F., Sempe, M., Guilloud-Bataille, M., and Patois, E. (1984). Adiposity rebound in children: a simple indicator for predicting obesity. Am J Clin Nutr 39, 129–135. 10.1093/ajcn/39.1.129.

4. Yengo, L., Sidorenko, J., Kemper, K.E., Zheng, Z., Wood, A.R., Weedon, M.N., Frayling, T.M., Hirschhorn, J., Yang, J., Visscher, P.M., and Consortium, G. (2018). Meta-analysis of genome-wide association studies for height and body mass index in approximately 700000 individuals of European ancestry. Hum Mol Genet 27, 3641–3649. 10.1093/hmg/ddy271.

5. Robinson, M.R., English, G., Moser, G., Lloyd-Jones, L.R., Triplett, M.A., Zhu, Z., Nolte, I.M., van Vliet-Ostaptchouk, J.V., Snieder, H., LifeLines Cohort, S., et al. (2017). Genotype-covariate interaction effects and the heritability of adult body mass index. Nat Genet 49, 1174–1181. 10.1038/ng.3912.

6. Elks, C.E., den Hoed, M., Zhao, J.H., Sharp, S.J., Wareham, N.J., Loos, R.J., and Ong, K.K. (2012). Variability in the heritability of body mass index: a systematic review and meta-regression. Front Endocrinol (Lausanne) 3, 29. 10.3389/fendo.2012.00029.

7. Kemper, K.E., Yengo, L., Zheng, Z., Abdellaoui, A., Keller, M.C., Goddard, M.E., Wray, N.R., Yang, J., and Visscher, P.M. (2021). Phenotypic covariance across the entire spectrum of relatedness for 86 billion pairs of individuals. Nat Commun 12, 1050. 10.1038/s41467-021-21283-4.

8. Silventoinen, K., Jelenkovic, A., Sund, R., Hur, Y.M., Yokoyama, Y., Honda, C., Hjelmborg, J., Moller, S., Ooki, S., Aaltonen, S., et al. (2016). Genetic and environmental effects on body mass index from infancy to the onset of adulthood: an individual-based pooled analysis of 45 twin cohorts participating in the Collaborative project of Development of Anthropometrical measures in Twins (CODATwins) study. Am J Clin Nutr 104, 371–379. 10.3945/ajcn.116.130252.

9. Llewellyn, C.H., Trzaskowski, M., Plomin, R., and Wardle, J. (2014). From modeling to measurement: developmental trends in genetic influence on adiposity in childhood. Obesity (Silver Spring) 22, 1756–1761. 10.1002/oby.20756.

10. Haworth, C.M., Carnell, S., Meaburn, E.L., Davis, O.S., Plomin, R., and Wardle, J. (2008). Increasing heritability of BMI and stronger associations with the FTO gene over childhood. Obesity (Silver Spring) 16, 2663–2668. 10.1038/oby.2008.434.

11. Helgeland, O., Vaudel, M., Juliusson, P.B., Lingaas Holmen, O., Juodakis, J., Bacelis, J., Jacobsson, B., Lindekleiv, H., Hveem, K., Lie, R.T., et al. (2019). Genome-wide association study reveals dynamic role of genetic variation in infant and early childhood growth. Nat Commun 10, 4448. 10.1038/s41467-019-12308-0.

12. Helgeland, O., Vaudel, M., Sole-Navais, P., Flatley, C., Juodakis, J., Bacelis, J., Koloen, I.L., Knudsen, G.P., Johansson, B.B., Magnus, P., et al. (2022). Characterization of the genetic architecture of infant and early childhood body mass index. Nat Metab 4, 344–358. 10.1038/s42255-022-00549-1.

13. Silventoinen, K., Li, W., Jelenkovic, A., Sund, R., Yokoyama, Y., Aaltonen, S., Piirtola, M., Sugawara, M., Tanaka, M., Matsumoto, S., et al. (2022). Changing genetic architecture of body mass index from infancy to early adulthood: an individual based pooled analysis of 25 twin cohorts. Int J Obes (Lond) 46, 1901–1909. 10.1038/s41366-022-01202-3.

14. Couto Alves, A., De Silva, N.M.G., Karhunen, V., Sovio, U., Das, S., Taal, H.R., Warrington, N.M., Lewin, A.M., Kaakinen, M., Cousminer, D.L., et al. (2019). GWAS on longitudinal growth traits reveals different genetic factors influencing infant, child, and adult BMI. Sci Adv 5, eaaw3095. 10.1126/sciadv.aaw3095.

15. Henderson, C.R., Jr. (1982). Analysis of covariance in the mixed model: higher-level, nonhomogeneous, and random regressions. Biometrics 38, 623–640.

16. Laird, N.M., and Ware, J.H. (1982). Random-effects models for longitudinal data. Biometrics 38, 963–974.

17. Yang, J., Lee, S.H., Goddard, M.E., and Visscher, P.M. (2011). GCTA: a tool for genome-wide complex trait analysis. Am J Hum Genet 88, 76–82. 10.1016/j.ajhg.2010.11.011.

18. Boyd, A., Golding, J., Macleod, J., Lawlor, D.A., Fraser, A., Henderson, J., Molloy, L., Ness, A., Ring, S., and Davey Smith, G. (2013). Cohort Profile: the ‘children of the 90s’--the index offspring of the Avon Longitudinal Study of Parents and Children. Int J Epidemiol 42, 111–127. 10.1093/ije/dys064.

19. Fraser, A., Macdonald-Wallis, C., Tilling, K., Boyd, A., Golding, J., Davey Smith, G., Henderson, J., Macleod, J., Molloy, L., Ness, A., et al. (2013). Cohort Profile: the Avon Longitudinal Study of Parents and Children: ALSPAC mothers cohort. Int J Epidemiol 42, 97–110. 10.1093/ije/dys066.

20. ALSPAC (2024). http://www.bristol.ac.uk/alspac/researchers/our-data/.

21. 1000 Genomes Project Consortiums, Auton, A., Brooks, L.D., Durbin, R.M., Garrison, E.P., Kang, H.M., Korbel, J.O., Marchini, J.L., McCarthy, S., McVean, G.A., and Abecasis, G.R. (2015). A global reference for human genetic variation. Nature 526, 68–74. 10.1038/nature15393.

22. Howie, B.N., Donnelly, P., and Marchini, J. (2009). A flexible and accurate genotype imputation method for the next generation of genome-wide association studies. PLoS Genet 5, e1000529. 10.1371/journal.pgen.1000529.

23. Howie, B., Marchini, J., and Stephens, M. (2011). Genotype imputation with thousands of genomes. G3 (Bethesda) 1, 457–470. 10.1534/g3.111.001198.

24. International HapMap, C. (2003). The International HapMap Project. Nature 426, 789–796. 10.1038/nature02168.

25. Daymont, C., Ross, M.E., Russell Localio, A., Fiks, A.G., Wasserman, R.C., and Grundmeier, R.W. (2017). Automated identification of implausible values in growth data from pediatric electronic health records. J Am Med Inform Assoc 24, 1080–1087. 10.1093/jamia/ocx037.

26. R Core Team (2022). R: A Language and Environment for Statistical Computing.

27. Warrington, N.M., Wu, Y.Y., Pennell, C.E., Marsh, J.A., Beilin, L.J., Palmer, L.J., Lye, S.J., and Briollais, L. (2013). Modelling BMI trajectories in children for genetic association studies. PLoS One 8, e53897. 10.1371/journal.pone.0053897.

28. Kirkpatrick, M., Lofsvold, D., and Bulmer, M. (1990). Analysis of the inheritance, selection and evolution of growth trajectories. Genetics 124, 979–993. 10.1093/genetics/124.4.979.

29. Mrode, R.A., and Thompson, R. (2014). Linear Models for the Prediction of Animal Breeding Values (CABI).

30. Fischer, T.M., Gilmour, A.R., and van der Werf, J.H. (2004). Computing approximate standard errors for genetic parameters derived from random regression models fitted by average information REML. Genet Sel Evol 36, 363–369. 10.1186/1297-9686-36-3-363.

31. Kirkpatrick, M., and Heckman, N. (1989). A quantitative genetic model for growth, shape, reaction norms, and other infinite-dimensional characters. J Math Biol 27, 429–450. 10.1007/BF00290638.

32. Zheng, Z., Liu, S., Sidorenko, J., Wang, Y., Lin, T., Yengo, L., Turley, P., Ani, A., Wang, R., Nolte, I.M., et al. (2024). Leveraging functional genomic annotations and genome coverage to improve polygenic prediction of complex traits within and between ancestries. Nat Genet 56, 767–777. 10.1038/s41588-024-01704-y.

33. Bycroft, C., Freeman, C., Petkova, D., Band, G., Elliott, L.T., Sharp, K., Motyer, A., Vukcevic, D., Delaneau, O., O’Connell, J., et al. (2018). The UK Biobank resource with deep phenotyping and genomic data. Nature 562, 203–209. 10.1038/s41586-018-0579-z.

34. Sudlow, C., Gallacher, J., Allen, N., Beral, V., Burton, P., Danesh, J., Downey, P., Elliott, P., Green, J., Landray, M., et al. (2015). UK biobank: an open access resource for identifying the causes of a wide range of complex diseases of middle and old age. PLoS Med 12, e1001779. 10.1371/journal.pmed.1001779.

35. Purcell, S., Neale, B., Todd-Brown, K., Thomas, L., Ferreira, M.A., Bender, D., Maller, J., Sklar, P., de Bakker, P.I., Daly, M.J., and Sham, P.C. (2007). PLINK: a tool set for whole-genome association and population-based linkage analyses. Am J Hum Genet 81, 559–575. 10.1086/519795.

36. Kuczmarski, R.J., Ogden, C.L., Guo, S.S., Grummer-Strawn, L.M., Flegal, K.M., Mei, Z., Wei, R., Curtin, L.R., Roche, A.F., and Johnson, C.L. (2002). 2000 CDC Growth Charts for the United States: methods and development. Vital Health Stat 11, 1–190.

37. Geserick, M., Vogel, M., Gausche, R., Lipek, T., Spielau, U., Keller, E., Pfaffle, R., Kiess, W., and Korner, A. (2018). Acceleration of BMI in Early Childhood and Risk of Sustained Obesity. N Engl J Med 379, 1303–1312. 10.1056/NEJMoa1803527.

38. Waters, D.L., Clark, S.A., Moghaddar, N., and van der Werf, J.H. (2022). Genomic analysis of the slope of the reaction norm for body weight in Australian sheep. Genet Sel Evol 54, 40. 10.1186/s12711-022-00734-6.

