## Supplementary Materials for "Distinct genetic profiles influence body mass index between infancy and adolescence"

**Supplementary Figures**

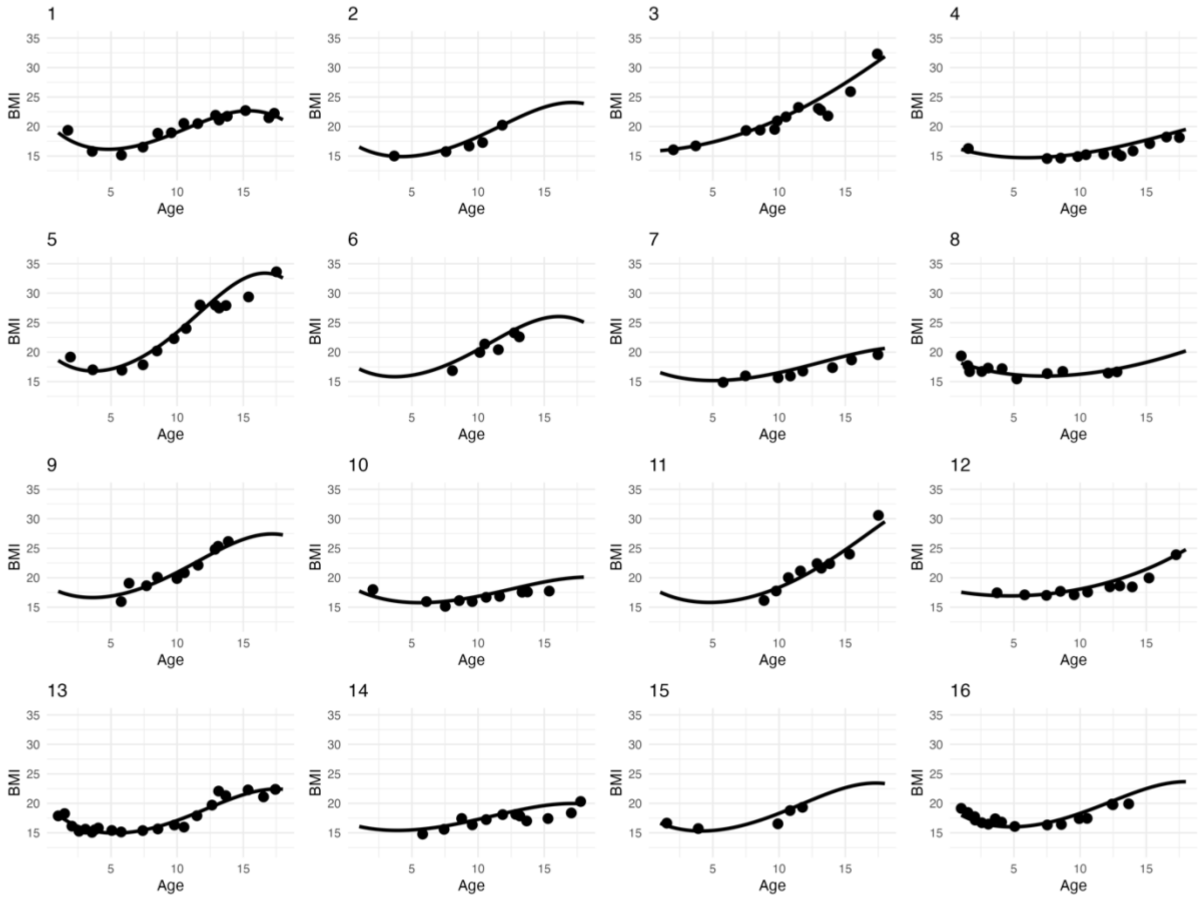

**Supplementary Figure 1: The fitted slopes (black line) of 16 randomly selected individuals with their observed BMI measurements from 1 to 18 years (black dots).**

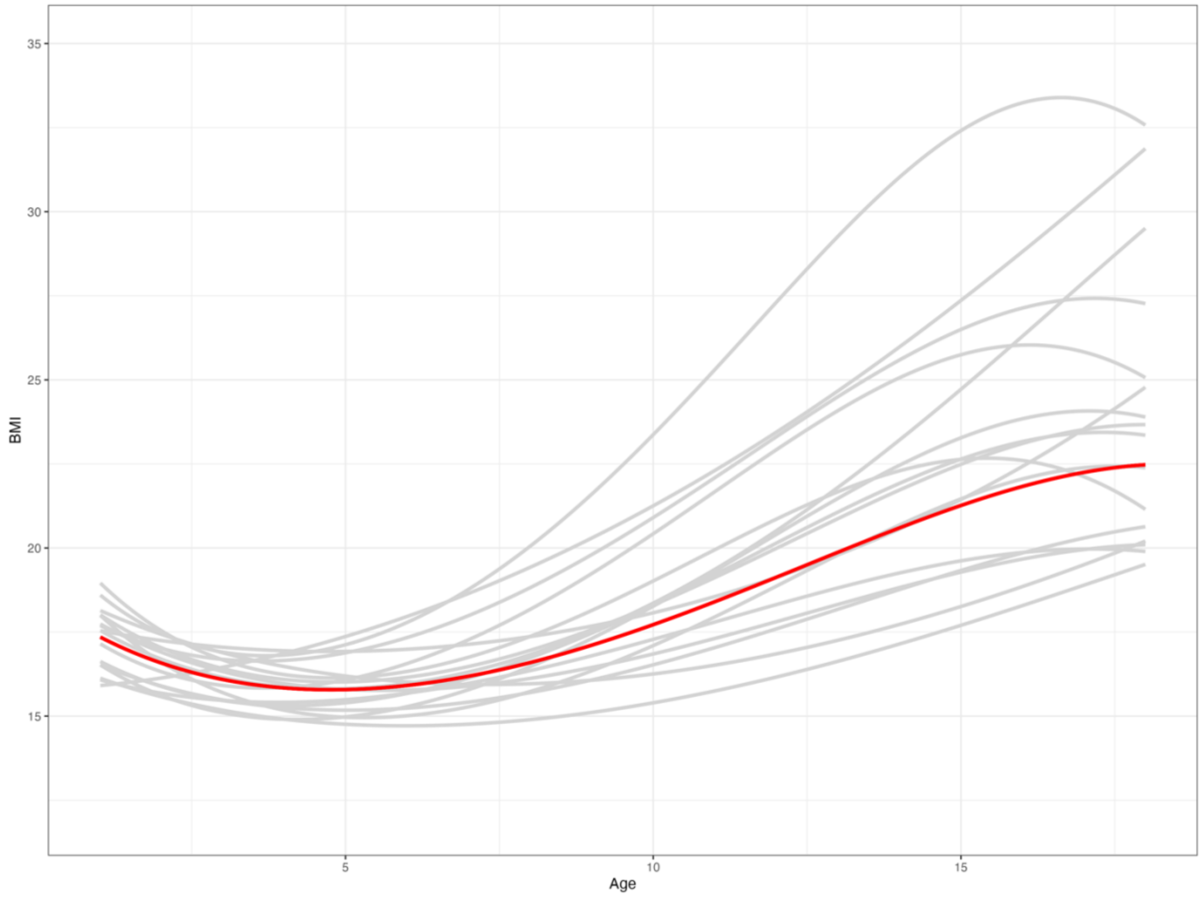

**Supplementary Figure 2: The fitted slopes of 16 randomly selected individuals from 1 to 18 years (grey lines) and the population mean (red line).**

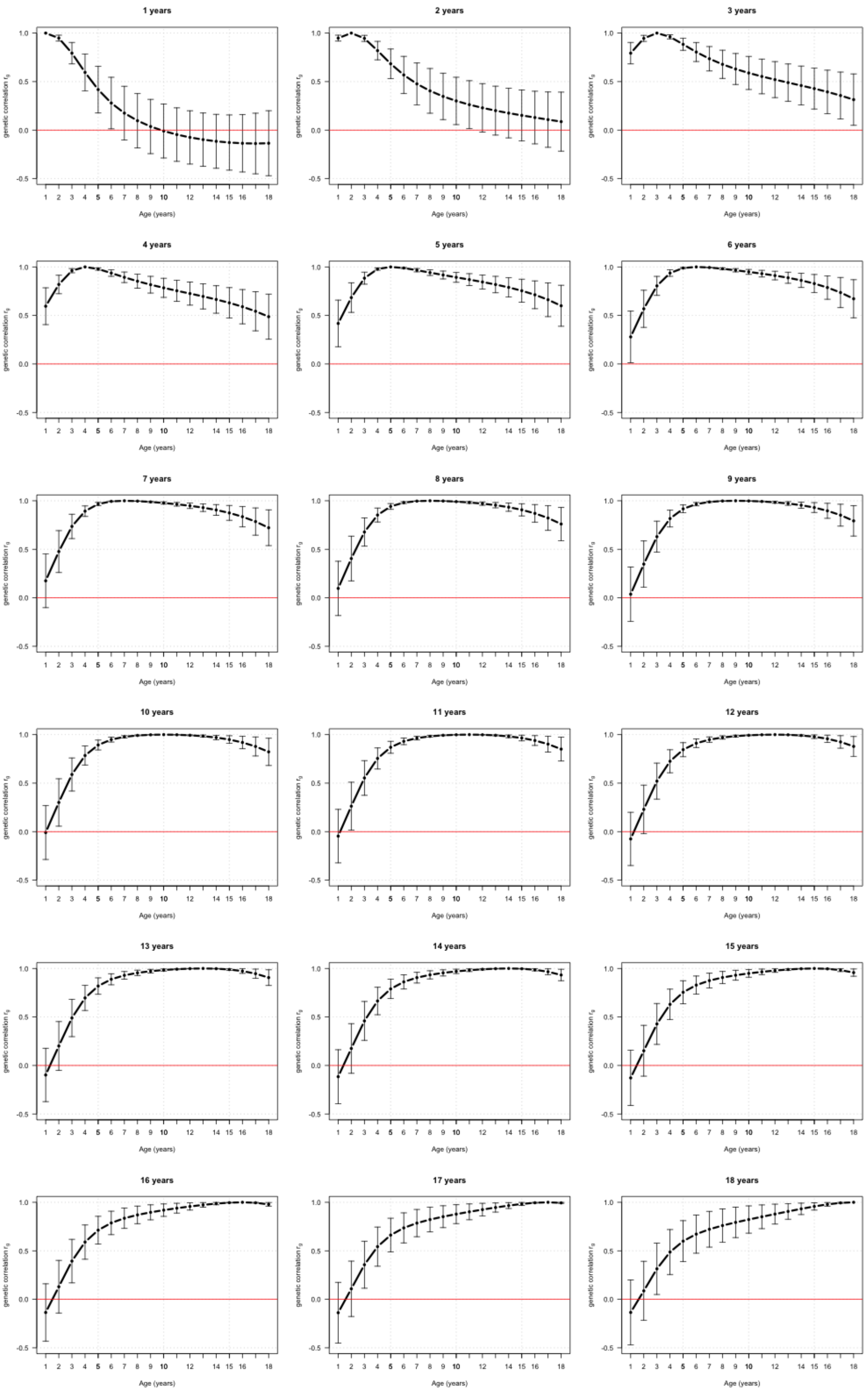

**Supplementary Figure 3: Estimates of genetic correlation ($r_g$) of BMI between different ages.**

Each plot represents the genetic correlations (represented along the y-axis) for a single age (given in the plot title) with all ages between 1 to 18 years in 1-year intervals (represented along the x-axis). Error bars represent the 95% confidence intervals. The red horizontal line indicates when genetic correlation is zero.

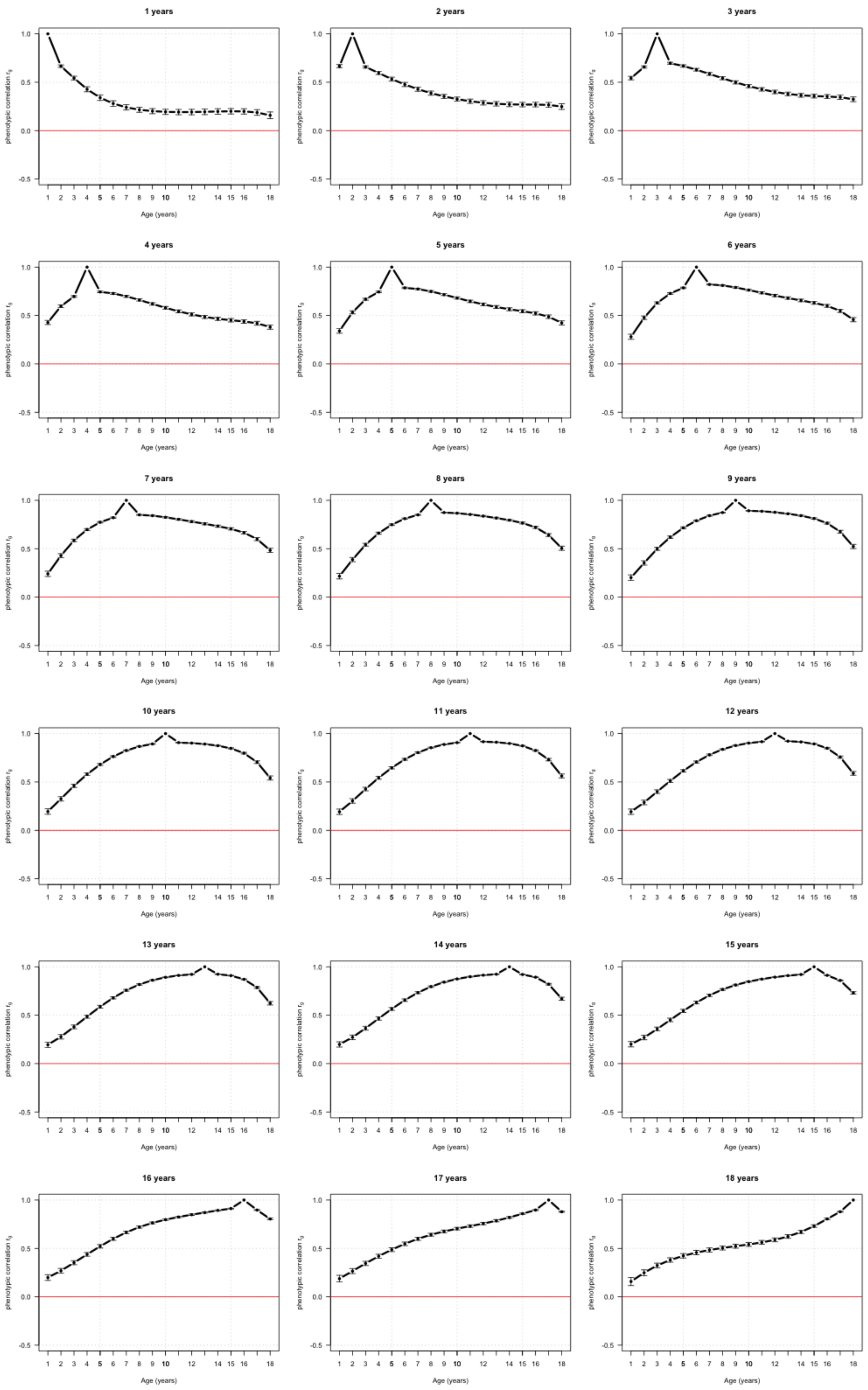
1 years
2 years
3 years
4 years
5 years
6 years
7 years
8 years
9 years
10 years
11 years
12 years
13 years
14 years
15 years
16 years
17 years
18 years
phenotypic correlation r_g
Age (years)

**Supplementary Figure 4: Estimates of phenotypic correlation (*$r_p$*) of BMI between different ages.**

Each plot represents the phenotypic correlations (y-axis) for a single age (given in the plot title) with all ages between 1 to 18 years in 1-year intervals (x-axis). Error bars represent the 95% confidence intervals. The red horizontal line indicates when phenotypic is zero.

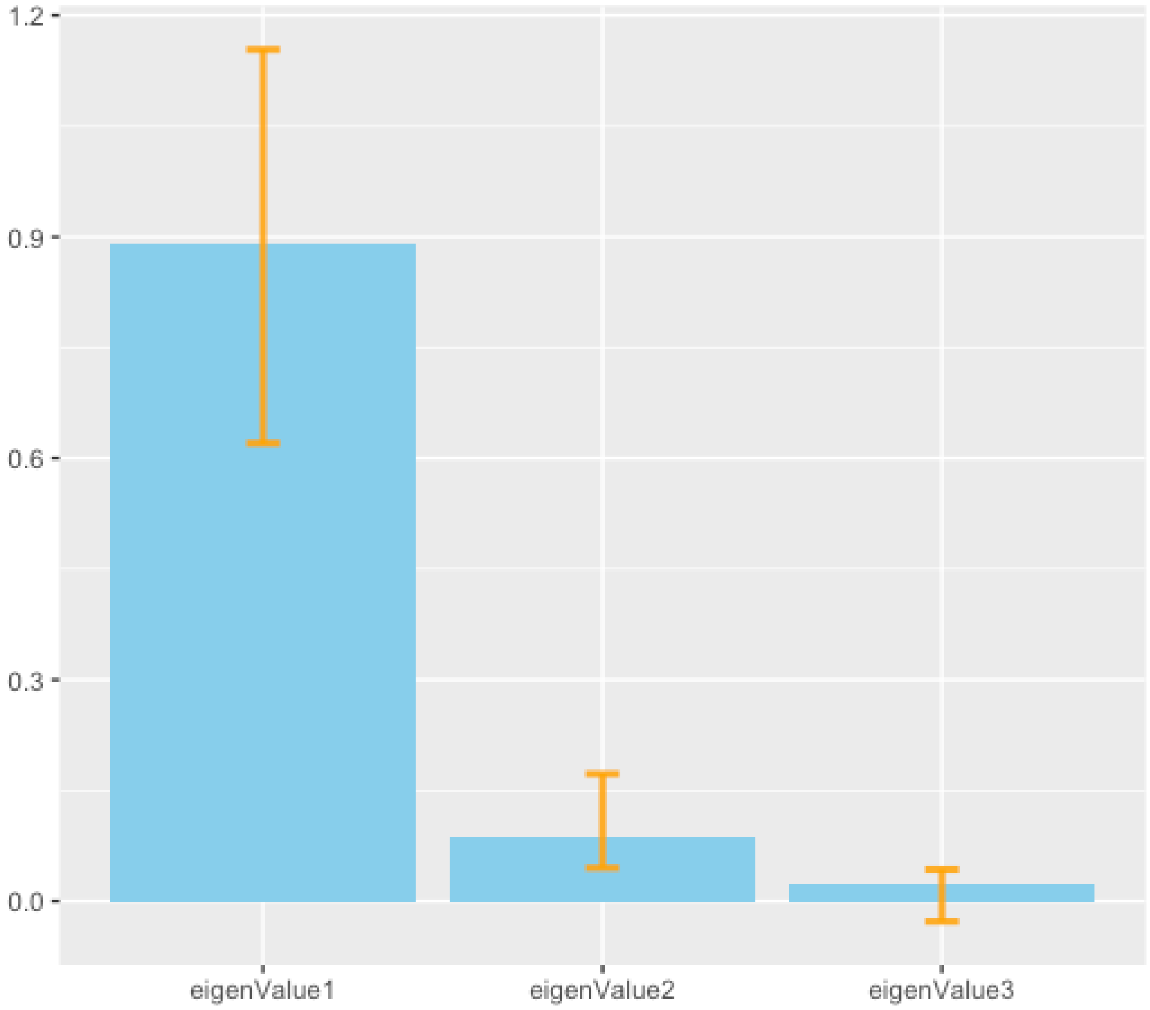

**Supplementary Figure 5**: **Proportion of the total variance explained by eigenvalues 1-3 after eigenvalue decomposition of the genetic variance-covariance matrix ($K_g$).**
The orange error bars are 95% confidence intervals calculated by numerical simulation.

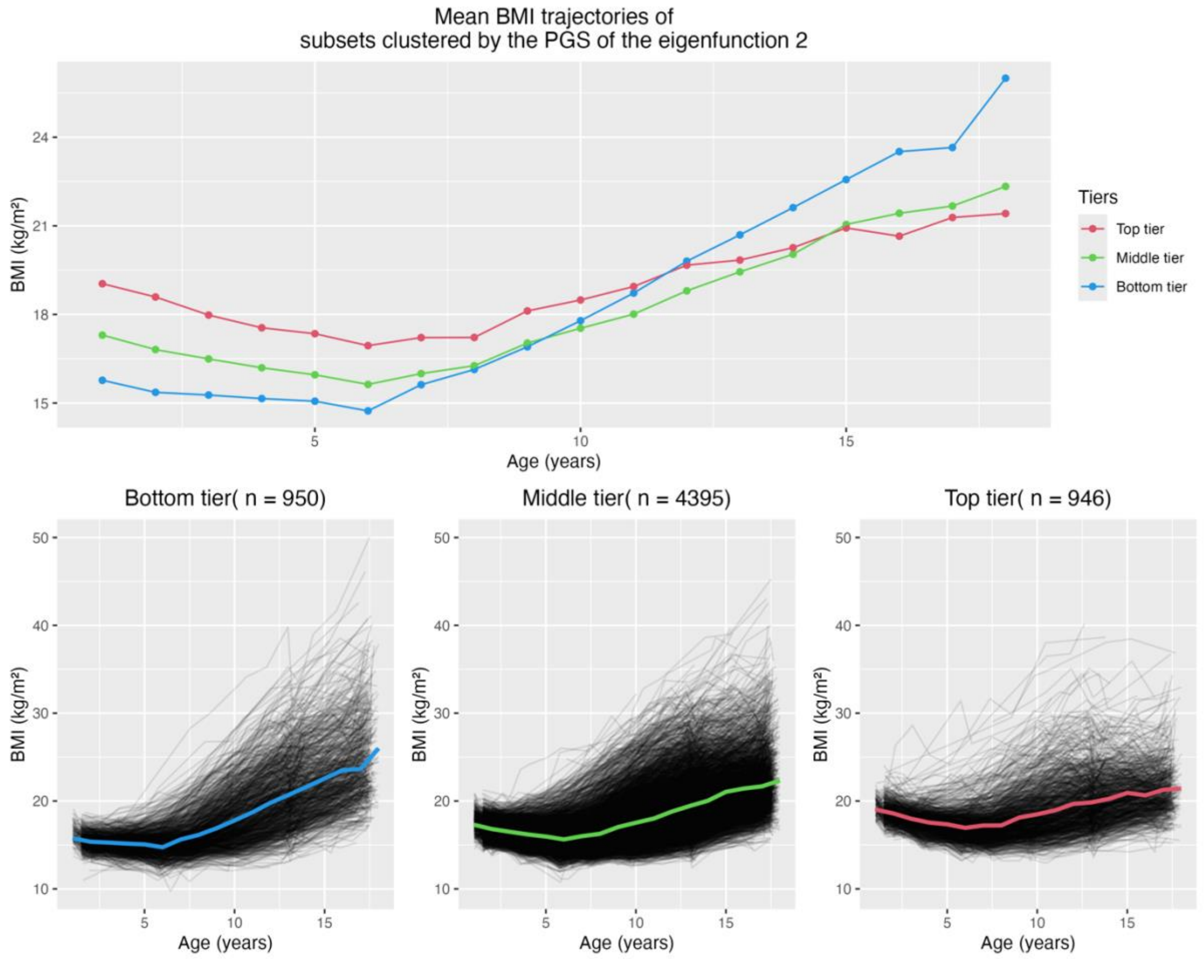

Supplementary Figure **6: Mean body mass index (BMI) from one to 18 years of age for three clusters based on a polygenic score (PGS) of eigenfunction two.**

The upper plot illustrates the mean BMI of each of the three clusters classified by PGS of principal components 2 (middle tier [individuals within one standard deviation of the mean PGS], top tier, and bottom tier of the residual) across yearly age bins ranging from 1 to 18 years. The lower plots separately display the mean BMI of each yearly age bin (colored lines) and individual trajectories for these three clusters (black lines).

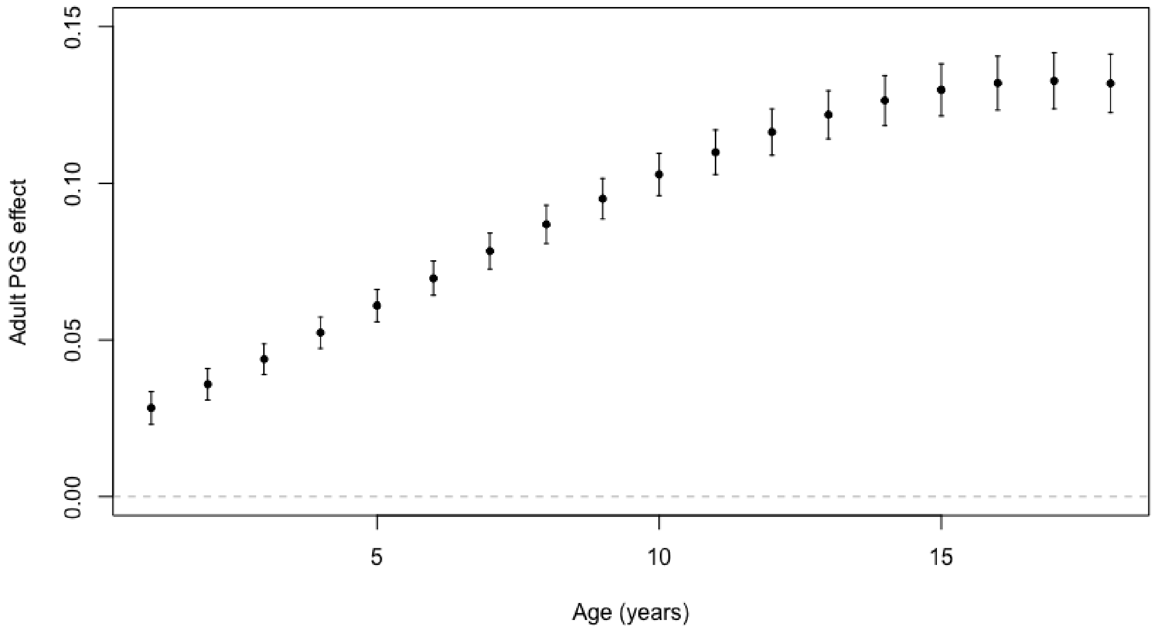

**Supplementary Figure 7: Estimated fixed effects of the polygenic score (PGS) of adult BMI on log(BMI) at different ages.**

The error bars are 95% confidence intervals calculated by the ASREML output of the prediction variance matrix (.vrb file). The mean log-transformed BMI in the studied cohort is 2.89, with a standard deviation of 0.17.

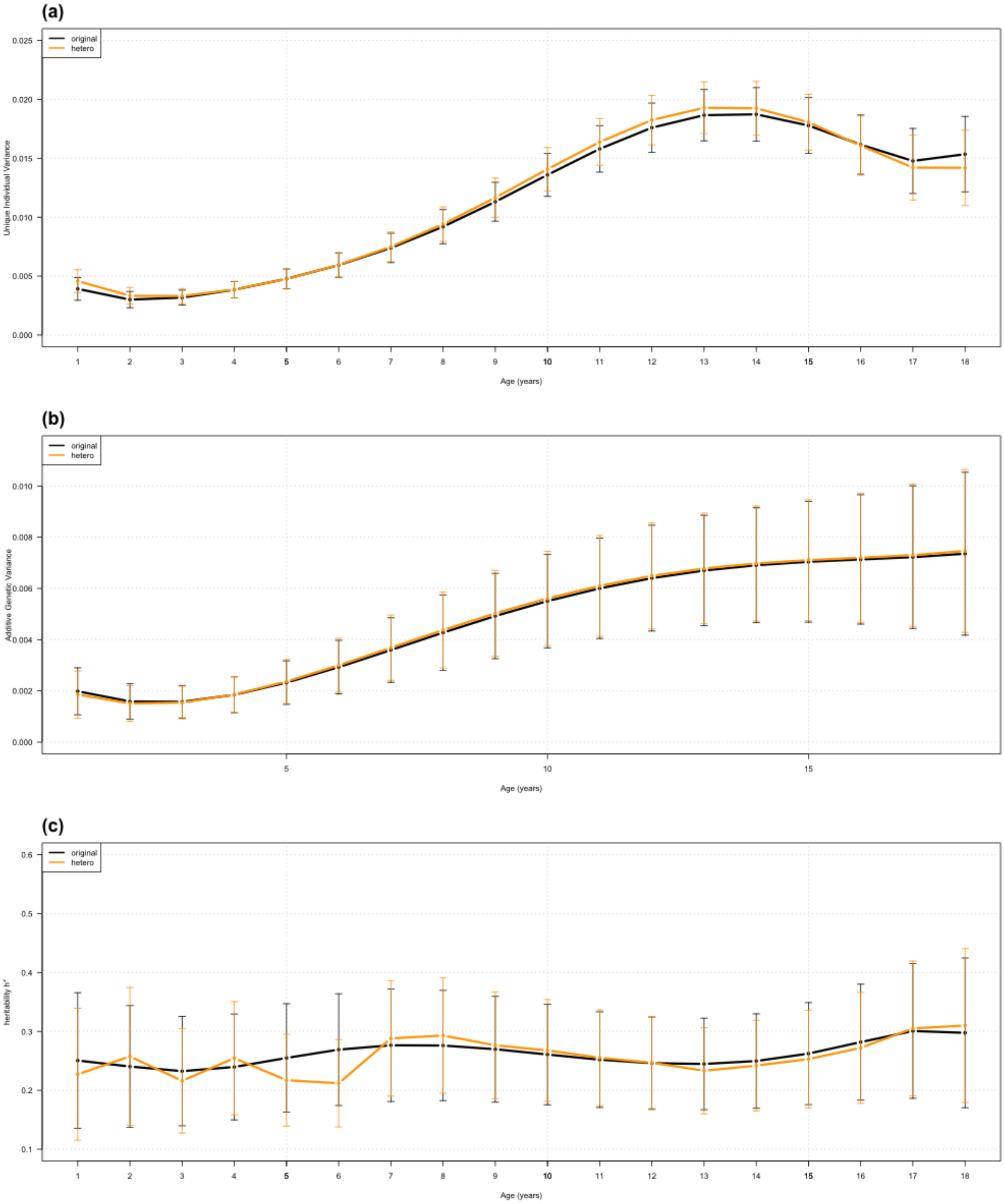

Supplementary Figure **8: Unique individual variances, additive genetic variances, and SNP-based heritability between homogeneous and heterogeneous error variance models**

The upper plot (a) presents the change of variances of unique individual effects as age increases using homogeneous (black) and heterogeneous (yellow) error variance models; the middle plot (b) presents the change of variances of additive genetic effects as age increases using homogeneous (black) and heterogeneous (yellow) error variance models; the lower plot (c) shows the estimated SNP-based heritability of BMI from 1 to 18 years of age using homogeneous (black) and heterogeneous (yellow) error variance models. Original: homogeneous error variance model; hetero: heterogeneous error variance model.

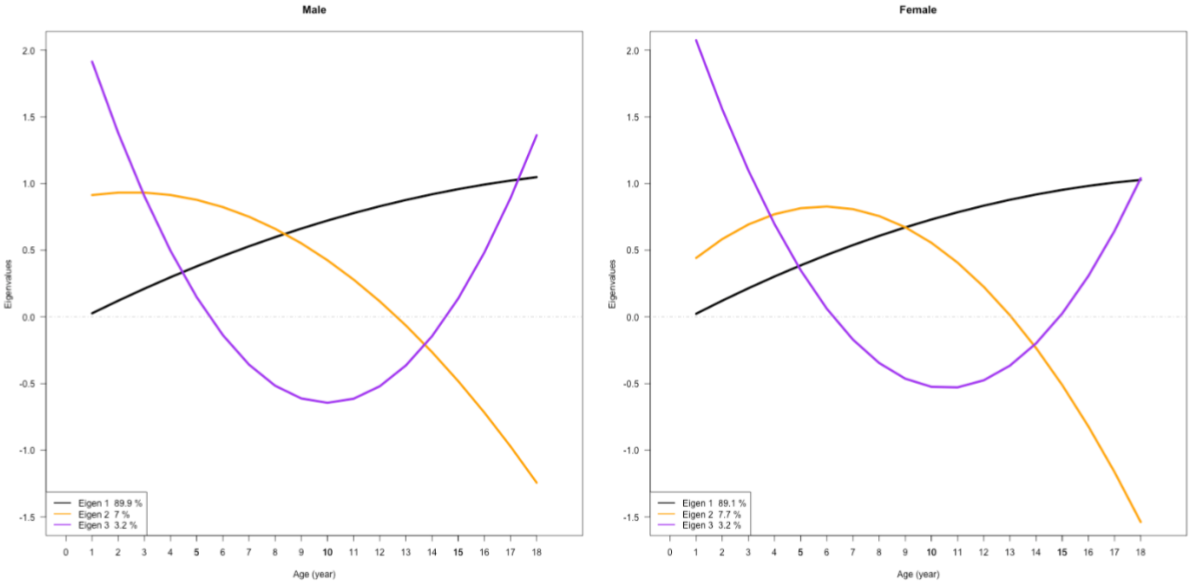

**Supplementary Figure 9: Eigenfunctions of additive genetic effects on BMI for male and female twins in CODATwins project evaluated from 1 to 18 years of age**

This figure illustrates the eigenfunctions representing the additive genetic effects on BMI as age changes in male (left) and female (right) twins in the CODATwins project. The x-axis denotes age in years, while the y-axis represents eigenvalues. Eigenfunctions 1, 2, and 3 are represented in lines in different colours (black, orange and purple, respectively). Each depicts the changing genetic effects of different eigens on BMI across different ages.

**Supplementary Tables**

**Supplementary Table 1: Statistics for the models with different order of polynomials in random effect terms for additive genetics.**

| Order of polynomial | n | AIC | LogL | P |
|---|---|---|---|---|
| **Intercept** | 12 | -293986.62 | 147005.31 | - |
| **Intercept, age** | 14 | -294012.80 | 147020.40 | $8.80 \times 10^{-5}$ |
| **Intercept, age, age$^2$,** | 17 | -294024.55 | 147029.27 | $4.97 \times 10^{-4}$ |
| **Intercept, age, age$^2$, age$^3$** | 21 | -294016.61 | 147029.31 | 0.999 |

LogL: log-likelihood; P: P-value of likelihood Number of parameters estimated (n), Alkaline Information Criterion (AIC) and log-likeihood (LogL) for the models with different orders of polynomials for additive genetics effects in the random regression model. Also shown are P-values from log-likelihood ratio tests between models with n vs. n-1 order polynomials.

**Supplementary Table 2: Estimates of fixed effects from the random regression model.**

| Model Term | Level | Fixed effects | SE | *P value* |
|---|---|---|---|---|
| Intercept | - | 2.9020 | 0.0019 | 0 |
| Legendre polynomials | slope | 0.1619 | 0.0014 | 0 |
| | $slope^2$ | 0.0535 | 0.0009 | 0 |
| | $slope^3$ | -0.0344 | 0.0007 | 0 |
| Source(clinic/questionnaire) | questionnaire | -0.0171 | 0.0006 | 0 |
| Sex(male/female) | female | -0.0111 | 0.0028 | 6.94E-05 |
| Interaction between sex and Legendre polynomials | female*slope | -0.0207 | 0.0020 | 0 |
| | female*$slope^2$ | 0.0093 | 0.0013 | 4.58E-12 |
| | female*$slope^3$ | 0.0044 | 0.0009 | 3.31E-06 |

Source: measurement sources; SE: standard errors of estimated fixed effects. * P value = 0 stands for < $2.225\times10^{-308}$.

**Supplementary Table 3: Variance-covariance matrices for random effect terms in the random regression model.**

| | | | | |
|---|---|---|---|---|
| **Unique Individual Variance-Covariance Matrix ($K_i$)** | | | | |
| | intercept | slope | $slope^2$ | $slope^3$ |
| intercept | 0.0165 (0.0012) | | | |
| slope | 0.0053 (0.0005) | 0.0035 (0.0003) | | |
| $slope^2$ | -0.0028 (0.0003) | -0.0006 (0.0001) | 0.0017 (0.0000) | |
| $slope^3$ | -0.0017 (0.0001) | -0.0010 (0.0000) | 0.0005 (0.0000) | 0.0008 (0.0000) |
| **Additive Genetic Variance-Covariance Matrix ($K_g$)** | | | | |
| | intercept | slope | $slope^2$ | |
| intercept | 0.0073 (0.0013) | | | |
| slope | 0.0024 (0.0005) | 0.0017 (0.0003) | | |
| $slope^2$ | -0.0012 (0.0003) | -0.0004 (0.0001) | 0.0004 (0.0001) | |
| **Residual Variance ($\sigma_e^2$)** | | | | |
| 0.0020 (0.0000) | | | | |

The variances (SE) are on the diagonals and the values below the diagonal are the covariances (SE)

**Supplementary Table 4**: **Estimated variance components and heritability of BMI at yearly intervals from one to 18 years of age from the random regression model**

| Age (years) | $V_g$ | Var($V_g$) | $V_i$ | Var($V_i$) | $V_P$ | Var($V_P$) | $\sigma_e^2$ | Var($\sigma_e^2$) | $h^2_{SNP}$ | SE($h^2_{SNP}$) |
|---|---|---|---|---|---|---|---|---|---|---|
| **1** | $1.98 \times 10^{-3}$ | $2.19 \times 10^{-7}$ | $3.91 \times 10^{-3}$ | $2.44 \times 10^{-7}$ | $7.92 \times 10^{-3}$ | $4.40 \times 10^{-8}$ | $2.02 \times 10^{-3}$ | $1.88 \times 10^{-10}$ | 0.251 | 0.059 |
| **2** | $1.59 \times 10^{-3}$ | $1.24 \times 10^{-7}$ | $3.00 \times 10^{-3}$ | $1.25 \times 10^{-7}$ | $6.61 \times 10^{-3}$ | $1.42 \times 10^{-8}$ | $2.02 \times 10^{-3}$ | $1.88 \times 10^{-10}$ | 0.240 | 0.053 |
| **3** | $1.57 \times 10^{-3}$ | $1.05 \times 10^{-7}$ | $3.18 \times 10^{-3}$ | $1.04 \times 10^{-7}$ | $6.77 \times 10^{-3}$ | $1.09 \times 10^{-8}$ | $2.02 \times 10^{-3}$ | $1.88 \times 10^{-10}$ | 0.233 | 0.047 |
| **4** | $1.85 \times 10^{-3}$ | $1.28 \times 10^{-7}$ | $3.84 \times 10^{-3}$ | $1.26 \times 10^{-7}$ | $7.71 \times 10^{-3}$ | $1.38 \times 10^{-8}$ | $2.02 \times 10^{-3}$ | $1.88 \times 10^{-10}$ | 0.240 | 0.046 |
| **5** | $2.32 \times 10^{-3}$ | $1.88 \times 10^{-7}$ | $4.77 \times 10^{-3}$ | $1.84 \times 10^{-7}$ | $9.11 \times 10^{-3}$ | $1.94 \times 10^{-8}$ | $2.02 \times 10^{-3}$ | $1.88 \times 10^{-10}$ | 0.255 | 0.047 |
| **6** | $2.93 \times 10^{-3}$ | $2.85 \times 10^{-7}$ | $5.93 \times 10^{-3}$ | $2.77 \times 10^{-7}$ | $1.09 \times 10^{-2}$ | $2.74 \times 10^{-8}$ | $2.02 \times 10^{-3}$ | $1.88 \times 10^{-10}$ | 0.269 | 0.048 |
| **7** | $3.60 \times 10^{-3}$ | $4.14 \times 10^{-7}$ | $7.39 \times 10^{-3}$ | $4.02 \times 10^{-7}$ | $1.30 \times 10^{-2}$ | $3.88 \times 10^{-8}$ | $2.02 \times 10^{-3}$ | $1.88 \times 10^{-10}$ | 0.276 | 0.049 |
| **8** | $4.27 \times 10^{-3}$ | $5.64 \times 10^{-7}$ | $9.19 \times 10^{-3}$ | $5.50 \times 10^{-7}$ | $1.55 \times 10^{-2}$ | $5.42 \times 10^{-8}$ | $2.02 \times 10^{-3}$ | $1.88 \times 10^{-10}$ | 0.276 | 0.048 |
| **9** | $4.92 \times 10^{-3}$ | $7.21 \times 10^{-7}$ | $1.13 \times 10^{-2}$ | $7.09 \times 10^{-7}$ | $1.82 \times 10^{-2}$ | $7.35 \times 10^{-8}$ | $2.02 \times 10^{-3}$ | $1.88 \times 10^{-10}$ | 0.270 | 0.046 |
| **10** | $5.51 \times 10^{-3}$ | $8.70 \times 10^{-7}$ | $1.36 \times 10^{-2}$ | $8.65 \times 10^{-7}$ | $2.11 \times 10^{-2}$ | $9.57 \times 10^{-8}$ | $2.02 \times 10^{-3}$ | $1.88 \times 10^{-10}$ | 0.261 | 0.044 |
| **11** | $6.00 \times 10^{-3}$ | $1.00 \times 10^{-6}$ | $1.58 \times 10^{-2}$ | $1.01 \times 10^{-6}$ | $2.38 \times 10^{-2}$ | $1.19 \times 10^{-7}$ | $2.02 \times 10^{-3}$ | $1.88 \times 10^{-10}$ | 0.252 | 0.041 |
| **12** | $6.40 \times 10^{-3}$ | $1.11 \times 10^{-6}$ | $1.76 \times 10^{-2}$ | $1.13 \times 10^{-6}$ | $2.60 \times 10^{-2}$ | $1.41 \times 10^{-7}$ | $2.02 \times 10^{-3}$ | $1.88 \times 10^{-10}$ | 0.246 | 0.040 |
| **13** | $6.70 \times 10^{-3}$ | $1.21 \times 10^{-6}$ | $1.87 \times 10^{-2}$ | $1.24 \times 10^{-6}$ | $2.74 \times 10^{-2}$ | $1.59 \times 10^{-7}$ | $2.02 \times 10^{-3}$ | $1.88 \times 10^{-10}$ | 0.245 | 0.040 |
| **14** | $6.91 \times 10^{-3}$ | $1.31 \times 10^{-6}$ | $1.87 \times 10^{-2}$ | $1.34 \times 10^{-6}$ | $2.77 \times 10^{-2}$ | $1.67 \times 10^{-7}$ | $2.02 \times 10^{-3}$ | $1.88 \times 10^{-10}$ | 0.250 | 0.041 |
| **15** | $7.04 \times 10^{-3}$ | $1.45 \times 10^{-6}$ | $1.78 \times 10^{-2}$ | $1.46 \times 10^{-6}$ | $2.68 \times 10^{-2}$ | $1.63 \times 10^{-7}$ | $2.02 \times 10^{-3}$ | $1.88 \times 10^{-10}$ | 0.262 | 0.044 |
| **16** | $7.13 \times 10^{-3}$ | $1.66 \times 10^{-6}$ | $1.62 \times 10^{-2}$ | $1.64 \times 10^{-6}$ | $2.53 \times 10^{-2}$ | $1.54 \times 10^{-7}$ | $2.02 \times 10^{-3}$ | $1.88 \times 10^{-10}$ | 0.282 | 0.050 |
| **17** | $7.22 \times 10^{-3}$ | $2.02 \times 10^{-6}$ | $1.48 \times 10^{-2}$ | $1.98 \times 10^{-6}$ | $2.40 \times 10^{-2}$ | $1.67 \times 10^{-7}$ | $2.02 \times 10^{-3}$ | $1.88 \times 10^{-10}$ | 0.301 | 0.058 |
| **18** | $7.36 \times 10^{-3}$ | $2.63 \times 10^{-6}$ | $1.53 \times 10^{-2}$ | $2.67 \times 10^{-6}$ | $2.47 \times 10^{-2}$ | $2.85 \times 10^{-7}$ | $2.02 \times 10^{-3}$ | $1.88 \times 10^{-10}$ | 0.298 | 0.065 |

$V_g$: estimated additive genetic variance; Var($V_g$): estimated variance of the additive genetic variance component ($V_g$); $V_i$: estimated unique individual variance; Var($V_i$): estimated variance of unique individual variance component; $V_P$: estimated phenotypic variance; Var($V_P$): estimated variance of phenotypic variance; $\sigma_e^2$: estimated residual variance; Var($\sigma_e^2$): estimated variance of residual variance component. $h^2_{snp}$: estimated SNP-heritability; SE($h^2_{snp}$): standard error of SNP-heritability estimate.

**Supplementary Table 5**: **Estimated genetic correlations between BMI at yearly intervals from one to 18 years from the random regression model.**

| Age (years) | 1 | 2 | 3 | 4 | 5 | 6 | 7 | 8 | 9 | 10 | 11 | 12 | 13 | 14 | 15 | 16 | 17 | 18 |
|---|---|---|---|---|---|---|---|---|---|---|---|---|---|---|---|---|---|---|
| **1** | 1 | | | | | | | | | | | | | | | | | |
| **2** | 0.948 (0.015) | 1 | | | | | | | | | | | | | | | | |
| **3** | 0.793 (0.056) | 0.945 (0.016) | 1 | | | | | | | | | | | | | | | |
| **4** | 0.595 (0.097) | 0.819 (0.049) | 0.961 (0.011) | 1 | | | | | | | | | | | | | | |
| **5** | 0.418 (0.123) | 0.684 (0.078) | 0.884 (0.031) | 0.979 (0.006) | 1 | | | | | | | | | | | | | |
| **6** | 0.279 (0.136) | 0.568 (0.098) | 0.805 (0.05) | 0.937 (0.017) | 0.989 (0.003) | 1 | | | | | | | | | | | | |
| **7** | 0.175 (0.141) | 0.477 (0.11) | 0.736 (0.064) | 0.893 (0.028) | 0.966 (0.009) | 0.994 (0.002) | 1 | | | | | | | | | | | |
| **8** | 0.097 (0.143) | 0.405 (0.118) | 0.678 (0.074) | 0.853 (0.037) | 0.942 (0.015) | 0.981 (0.005) | 0.996 (0.001) | 1 | | | | | | | | | | |
| **9** | 0.037 (0.143) | 0.347 (0.122) | 0.63 (0.082) | 0.817 (0.044) | 0.917 (0.021) | 0.966 (0.009) | 0.988 (0.003) | 0.998 (0.001) | 1 | | | | | | | | | |
| **10** | -0.009 (0.142) | 0.301 (0.125) | 0.589 (0.087) | 0.784 (0.05) | 0.893 (0.026) | 0.949 (0.013) | 0.977 (0.006) | 0.992 (0.002) | 0.998 (0.001) | 1 | | | | | | | | |
| **11** | -0.046 (0.141) | 0.262 (0.126) | 0.553 (0.091) | 0.754 (0.055) | 0.869 (0.031) | 0.931 (0.017) | 0.964 (0.01) | 0.983 (0.005) | 0.993 (0.002) | 0.998 (0.001) | 1 | | | | | | | |
| **12** | -0.075 (0.140) | 0.229 (0.127) | 0.521 (0.095) | 0.725 (0.061) | 0.845 (0.037) | 0.911 (0.023) | 0.948 (0.014) | 0.971 (0.009) | 0.984 (0.005) | 0.993 (0.002) | 0.998 (0.001) | 1 | | | | | | |
| **13** | -0.098 (0.140) | 0.201 (0.129) | 0.49 (0.098) | 0.696 (0.066) | 0.819 (0.043) | 0.888 (0.029) | 0.929 (0.02) | 0.955 (0.014) | 0.972 (0.009) | 0.984 (0.006) | 0.993 (0.003) | 0.998 (0.001) | 1 | | | | | |
| **14** | -0.115 (0.142) | 0.175 (0.130) | 0.459 (0.102) | 0.665 (0.072) | 0.789 (0.051) | 0.861 (0.037) | 0.905 (0.028) | 0.934 (0.022) | 0.954 (0.016) | 0.97 (0.011) | 0.982 (0.007) | 0.991 (0.004) | 0.998 (0.001) | 1 | | | | |
| **15** | -0.128 (0.145) | 0.152 (0.134) | 0.428 (0.107) | 0.630 (0.08) | 0.755 (0.061) | 0.829 (0.048) | 0.875 (0.039) | 0.907 (0.032) | 0.93 (0.026) | 0.949 (0.02) | 0.965 (0.015) | 0.978 (0.009) | 0.989 (0.005) | 0.997 (0.001) | 1 | | | |
| **16** | -0.136 (0.151) | 0.130 (0.138) | 0.394 (0.114) | 0.59 (0.09) | 0.713 (0.073) | 0.788 (0.062) | 0.836 (0.053) | 0.87 (0.046) | 0.897 (0.040) | 0.919 (0.033) | 0.939 (0.026) | 0.957 (0.019) | 0.973 (0.012) | 0.987 (0.006) | 0.996 (0.002) | 1 | | |
| **17** | -0.138 (0.159) | 0.108 (0.146) | 0.356 (0.123) | 0.543 (0.103) | 0.662 (0.089) | 0.736 (0.079) | 0.786 (0.072) | 0.822 (0.065) | 0.852 (0.058) | 0.878 (0.05) | 0.902 (0.042) | 0.925 (0.033) | 0.946 (0.024) | 0.966 (0.015) | 0.983 (0.008) | 0.995 (0.002) | 1 | |
| **18** | -0.135 (0.171) | 0.087 (0.155) | 0.314 (0.135) | 0.487 (0.118) | 0.600 (0.108) | 0.672 (0.100) | 0.722 (0.094) | 0.76 (0.087) | 0.793 (0.080) | 0.822 (0.072) | 0.851 (0.062) | 0.879 (0.052) | 0.906 (0.041) | 0.933 (0.030) | 0.957 (0.019) | 0.979 (0.010) | 0.994 (0.003) | 1 |

The lower triangle shows the genetic correlations between BMI across different ages (in years) as indicated in the column and row headers. Standard errors for each estimate are given in brackets.

**Supplementary Table 6**: **Estimated phenotypic correlations between BMI at yearly intervals from one to 18 years from the random regression model.**

| **Age (years)** | **1** | **2** | **3** | **4** | **5** | **6** | **7** | **8** | **9** | **10** | **11** | **12** | **13** | **14** | **15** | **16** | **17** | **18** |
|---|---|---|---|---|---|---|---|---|---|---|---|---|---|---|---|---|---|---|
| **1** | 1 | | | | | | | | | | | | | | | | | |
| **2** | 0.666 (0.008) | 1 | | | | | | | | | | | | | | | | |
| **3** | 0.543 (0.009) | 0.658 (0.006) | 1 | | | | | | | | | | | | | | | |
| **4** | 0.428 (0.011) | 0.596 (0.007) | 0.696 (0.005) | 1 | | | | | | | | | | | | | | |
| **5** | 0.34 (0.012) | 0.533 (0.008) | 0.668 (0.005) | 0.744 (0.004) | 1 | | | | | | | | | | | | | |
| **6** | 0.280 (0.013) | 0.477 (0.009) | 0.629 (0.006) | 0.728 (0.004) | 0.786 (0.003) | 1 | | | | | | | | | | | | |
| **7** | 0.241 (0.014) | 0.429 (0.01) | 0.586 (0.007) | 0.698 (0.005) | 0.774 (0.004) | 0.821 (0.003) | 1 | | | | | | | | | | | |
| **8** | 0.216 (0.014) | 0.388 (0.011) | 0.541 (0.008) | 0.660 (0.005) | 0.749 (0.004) | 0.811 (0.003) | 0.85 (0.003) | 1 | | | | | | | | | | |
| **9** | 0.202 (0.014) | 0.353 (0.011) | 0.499 (0.008) | 0.62 (0.006) | 0.716 (0.005) | 0.790 (0.004) | 0.842 (0.003) | 0.874 (0.002) | 1 | | | | | | | | | |
| **10** | 0.194 (0.014) | 0.325 (0.012) | 0.46 (0.009) | 0.579 (0.007) | 0.681 (0.005) | 0.763 (0.004) | 0.825 (0.003) | 0.867 (0.002) | 0.893 (0.002) | 1 | | | | | | | | |
| **11** | 0.192 (0.014) | 0.304 (0.012) | 0.427 (0.009) | 0.543 (0.007) | 0.646 (0.006) | 0.734 (0.005) | 0.804 (0.004) | 0.854 (0.003) | 0.887 (0.002) | 0.906 (0.002) | 1 | | | | | | | |
| **12** | 0.192 (0.014) | 0.288 (0.012) | 0.400 (0.010) | 0.511 (0.008) | 0.615 (0.007) | 0.706 (0.005) | 0.781 (0.004) | 0.837 (0.003) | 0.876 (0.002) | 0.902 (0.002) | 0.916 (0.001) | 1 | | | | | | |
| **13** | 0.195 (0.014) | 0.277 (0.012) | 0.379 (0.01) | 0.485 (0.008) | 0.588 (0.007) | 0.68 (0.006) | 0.757 (0.005) | 0.818 (0.003) | 0.861 (0.003) | 0.891 (0.002) | 0.911 (0.002) | 0.922 (0.001) | 1 | | | | | |
| **14** | 0.198 (0.014) | 0.271 (0.012) | 0.365 (0.010) | 0.466 (0.009) | 0.565 (0.008) | 0.655 (0.007) | 0.733 (0.005) | 0.795 (0.004) | 0.841 (0.003) | 0.875 (0.002) | 0.898 (0.002) | 0.914 (0.002) | 0.923 (0.001) | 1 | | | | |
| **15** | 0.200 (0.014) | 0.269 (0.012) | 0.357 (0.01) | 0.451 (0.009) | 0.544 (0.008) | 0.631 (0.007) | 0.705 (0.006) | 0.766 (0.005) | 0.812 (0.004) | 0.847 (0.003) | 0.873 (0.003) | 0.893 (0.002) | 0.909 (0.002) | 0.920 (0.002) | 1 | | | |
| **16** | 0.199 (0.015) | 0.269 (0.012) | 0.352 (0.01) | 0.438 (0.01) | 0.522 (0.009) | 0.599 (0.008) | 0.666 (0.007) | 0.721 (0.006) | 0.764 (0.005) | 0.797 (0.004) | 0.825 (0.004) | 0.849 (0.004) | 0.871 (0.003) | 0.893 (0.003) | 0.912 (0.002) | 1 | | |
| **17** | 0.188 (0.017) | 0.266 (0.013) | 0.345 (0.011) | 0.419 (0.010) | 0.487 (0.010) | 0.548 (0.009) | 0.599 (0.008) | 0.642 (0.007) | 0.676 (0.007) | 0.704 (0.007) | 0.73 (0.007) | 0.757 (0.006) | 0.786 (0.006) | 0.82 (0.005) | 0.859 (0.004) | 0.897 (0.002) | 1 | |
| **18** | 0.158 (0.021) | 0.249 (0.015) | 0.324 (0.012) | 0.380 (0.012) | 0.423 (0.012) | 0.457 (0.011) | 0.484 (0.011) | 0.505 (0.011) | 0.523 (0.010) | 0.541 (0.010) | 0.562 (0.010) | 0.588 (0.010) | 0.623 (0.009) | 0.670 (0.008) | 0.731 (0.007) | 0.805 (0.005) | 0.879 (0.003) | 1 |

The lower triangle shows the genetic correlations between BMI across different ages (in years) as indicated in the column and row headers. Standard errors for each estimate are given in brackets.

**Supplementary Table 7: Estimated fixed effects from the random regression model adjusting for adult BMI PGS.**

| *Model Term* | Level | Fixed effects | SE |
|---|---|---|---|
| Intercept | - | 2.8890 | 0.0019 |
| Legendre polynomials | slope | 0.1534 | 0.0013 |
| | $slope^2$ | 0.0560 | 0.0009 |
| | $slope^3$ | -0.0326 | 0.0007 |
| Source(clinic/questionnaire) | questionnaire | -0.0170 | 0.0006 |
| Sex(male/female) | female | -0.0119 | 0.0026 |
| Interaction terms between sex and Legendre polynomials | female*slope | -0.0211 | 0.0019 |
| | $female*slope^2$ | 0.0095 | 0.0013 |
| | $female*slope^3$ | 0.0046 | 0.0009 |
| PGS | - | 0.0990 | 0.0034 |
| Interaction terms between adult BMI and Legendre polynomials) | PGS*slope | 0.0657 | 0.0024 |
| | $PGS*slope^2$ | -0.0189 | 0.0017 |
| | $PGS*slope^3$ | -0.0140 | 0.0012 |

Source: measurement sources SE: standard errors of estimated fixed effect. PGS: polygenic score

**Supplementary Table 8 Estimated variances and covariances for random effect terms in the random regression model for adult BMI PGS.**

| | | | | |
|---|---|---|---|---|
| **Unique Individual Variance-Covariance Matrix($K_i$)** | | | | |
| | intercept | slope | slope$^2$ | slope$^3$ |
| intercept | 0.0164 (0.0011) | | | |
| slope | 0.0055 (0.0005) | 0.0037 (0.0003) | | |
| slope$^2$ | -0.0028 (0.0003) | -0.0006 (0.0001) | 0.0017 (0.0001) | |
| slope$^3$ | -0.0014 (0.0001) | -0.0008 (0.0000) | 0.0005 (0.0000) | 0.0007 (0.0000) |
| **Additive Genetic Variance-Covariance Matrix ($K_g$)** | | | | |
| | intercept | slope | slope$^2$ | |
| intercept | 0.0045 (0.0011) | | | |
| slope | 0.0008 (0.0004) | 0.0009 (0.0003) | | |
| slope$^2$ | -0.0008 (0.0003) | -0.0002 (0.0001) | 0.0004 (0.0001) | |
| **Residual Variance** | | | | |
| 0.0020 (0.0000) | | | | |

The variances (SE) are on the diagonals and the values below the diagonal are the covariances (SE)

**Supplementary Table 9**: **Estimated heritability of BMI at yearly intervals from one to 18 years of age from the random regression model conditioning on adult BMI PGS**

| Age (years) | $V_g$ | Var($V_g$) | $V_i$ | Var($V_i$) | $V_P$ | Var($V_P$) | $\sigma_e^2$ | Var($\sigma_e^2$) | $h^2_{SNP}$ | SE($h^2_{SNP}$) |
|---|---|---|---|---|---|---|---|---|---|---|
| **1** | $1.98 \times 10^{-3}$ | $2.19 \times 10^{-7}$ | $3.89 \times 10^{-3}$ | $2.40 \times 10^{-7}$ | $7.88 \times 10^{-3}$ | $4.40 \times 10^{-8}$ | $2.02 \times 10^{-3}$ | $1.88 \times 10^{-10}$ | 0.251 | 0.059 |
| **2** | $1.64 \times 10^{-3}$ | $1.23 \times 10^{-7}$ | $2.91 \times 10^{-3}$ | $1.23 \times 10^{-7}$ | $6.56 \times 10^{-3}$ | $1.42 \times 10^{-8}$ | $2.02 \times 10^{-3}$ | $1.88 \times 10^{-10}$ | 0.249 | 0.053 |
| **3** | $1.57 \times 10^{-3}$ | $1.00 \times 10^{-7}$ | $3.09 \times 10^{-3}$ | $9.89 \times 10^{-8}$ | $6.68 \times 10^{-3}$ | $1.09 \times 10^{-8}$ | $2.02 \times 10^{-3}$ | $1.88 \times 10^{-10}$ | 0.235 | 0.047 |
| **4** | $1.70 \times 10^{-3}$ | $1.16 \times 10^{-7}$ | $3.81 \times 10^{-3}$ | $1.16 \times 10^{-7}$ | $7.52 \times 10^{-3}$ | $1.38 \times 10^{-8}$ | $2.02 \times 10^{-3}$ | $1.88 \times 10^{-10}$ | 0.225 | 0.045 |
| **5** | $1.95 \times 10^{-3}$ | $1.64 \times 10^{-7}$ | $4.79 \times 10^{-3}$ | $1.64 \times 10^{-7}$ | $8.77 \times 10^{-3}$ | $1.94 \times 10^{-8}$ | $2.02 \times 10^{-3}$ | $1.88 \times 10^{-10}$ | 0.223 | 0.046 |
| **6** | $2.28 \times 10^{-3}$ | $2.41 \times 10^{-7}$ | $5.99 \times 10^{-3}$ | $2.41 \times 10^{-7}$ | $1.03 \times 10^{-2}$ | $2.74 \times 10^{-8}$ | $2.02 \times 10^{-3}$ | $1.88 \times 10^{-10}$ | 0.222 | 0.047 |
| **7** | $2.63 \times 10^{-3}$ | $3.43 \times 10^{-7}$ | $7.46 \times 10^{-3}$ | $3.42 \times 10^{-7}$ | $1.21 \times 10^{-2}$ | $3.88 \times 10^{-8}$ | $2.02 \times 10^{-3}$ | $1.88 \times 10^{-10}$ | 0.217 | 0.048 |
| **8** | $2.97 \times 10^{-3}$ | $4.58 \times 10^{-7}$ | $9.21 \times 10^{-3}$ | $4.61 \times 10^{-7}$ | $1.42 \times 10^{-2}$ | $5.42 \times 10^{-8}$ | $2.02 \times 10^{-3}$ | $1.88 \times 10^{-10}$ | 0.209 | 0.047 |
| **9** | $3.25 \times 10^{-3}$ | $5.74 \times 10^{-7}$ | $1.12 \times 10^{-2}$ | $5.84 \times 10^{-7}$ | $1.65 \times 10^{-2}$ | $7.35 \times 10^{-8}$ | $2.02 \times 10^{-3}$ | $1.88 \times 10^{-10}$ | 0.197 | 0.046 |
| **10** | $3.47 \times 10^{-3}$ | $6.78 \times 10^{-7}$ | $1.34 \times 10^{-2}$ | $7.01 \times 10^{-7}$ | $1.89 \times 10^{-2}$ | $9.57 \times 10^{-8}$ | $2.02 \times 10^{-3}$ | $1.88 \times 10^{-10}$ | 0.184 | 0.043 |
| **11** | $3.60 \times 10^{-3}$ | $7.65 \times 10^{-7}$ | $1.55 \times 10^{-2}$ | $8.03 \times 10^{-7}$ | $2.11 \times 10^{-2}$ | $1.19 \times 10^{-7}$ | $2.02 \times 10^{-3}$ | $1.88 \times 10^{-10}$ | 0.171 | 0.041 |
| **12** | $3.65 \times 10^{-3}$ | $8.32 \times 10^{-7}$ | $1.72 \times 10^{-2}$ | $8.88 \times 10^{-7}$ | $2.28 \times 10^{-2}$ | $1.41 \times 10^{-7}$ | $2.02 \times 10^{-3}$ | $1.88 \times 10^{-10}$ | 0.160 | 0.040 |
| **13** | $3.63 \times 10^{-3}$ | $8.88 \times 10^{-7}$ | $1.82 \times 10^{-2}$ | $9.59 \times 10^{-7}$ | $2.38 \times 10^{-2}$ | $1.59 \times 10^{-7}$ | $2.02 \times 10^{-3}$ | $1.88 \times 10^{-10}$ | 0.152 | 0.039 |
| **14** | $3.54 \times 10^{-3}$ | $9.49 \times 10^{-7}$ | $1.84 \times 10^{-2}$ | $1.03 \times 10^{-6}$ | $2.39 \times 10^{-2}$ | $1.67 \times 10^{-7}$ | $2.02 \times 10^{-3}$ | $1.88 \times 10^{-10}$ | 0.148 | 0.041 |
| **15** | $3.42 \times 10^{-3}$ | $1.04 \times 10^{-6}$ | $1.76 \times 10^{-2}$ | $1.11 \times 10^{-6}$ | $2.31 \times 10^{-2}$ | $1.63 \times 10^{-7}$ | $2.02 \times 10^{-3}$ | $1.88 \times 10^{-10}$ | 0.148 | 0.044 |
| **16** | $3.29 \times 10^{-3}$ | $1.20 \times 10^{-6}$ | $1.64 \times 10^{-2}$ | $1.26 \times 10^{-6}$ | $2.18 \times 10^{-2}$ | $1.54 \times 10^{-7}$ | $2.02 \times 10^{-3}$ | $1.88 \times 10^{-10}$ | 0.151 | 0.050 |
| **17** | $3.21 \times 10^{-3}$ | $1.48 \times 10^{-6}$ | $1.57 \times 10^{-2}$ | $1.55 \times 10^{-6}$ | $2.09 \times 10^{-2}$ | $1.67 \times 10^{-7}$ | $2.02 \times 10^{-3}$ | $1.88 \times 10^{-10}$ | 0.154 | 0.058 |
| **18** | $3.23 \times 10^{-3}$ | $1.99 \times 10^{-6}$ | $1.70 \times 10^{-2}$ | $2.16 \times 10^{-6}$ | $2.23 \times 10^{-2}$ | $2.85 \times 10^{-7}$ | $2.02 \times 10^{-3}$ | $1.88 \times 10^{-10}$ | 0.145 | 0.063 |

$V_g$: estimated additive genetic variance; Var($V_g$): estimated variance of the additive genetic variance component ($V_g$); $V_i$: estimated unique individual variance. Var($V_i$): estimated variance of unique individual variance component; $V_P$: estimated phenotypic variance; Var($V_P$): estimated variance of phenotypic variance; $\sigma_e^2$: estimated residual variance; Var($\sigma_e^2$): estimated variance of residual variance component. $h^2_{snp}$: estimated SNP-heritability; SE($h^2_{snp}$): standard error of SNP-heritability estimate.

**Supplementary Table 10**: **Estimated SNP-based heritability at multiple ages from cross-sectional genetic analyses in GCTA**

| Follow-up codes | Mean age (year) | $h^2_{SNP}$ | SE | P value | N |
|---|---|---|---|---|---|
| **Child health database 2** | 0.8 | 0.28 | 0.07 | $4.28 \times 10^{-6}$ | 4799 |
| **Child health database 3** | 1.7 | 0.26 | 0.07 | $1.13 \times 10^{-4}$ | 4454 |
| **Focus@7** | 7.6 | 0.26 | 0.06 | $6.20 \times 10^{-6}$ | 5350 |
| **Focus@10** | 10.7 | 0.28 | 0.06 | $5.33 \times 10^{-6}$ | 5228 |
| **Teen Focus2** | 13.9 | 0.23 | 0.07 | $7.81 \times 10^{-4}$ | 4404 |
| **Teen Focus3** | 15.5 | 0.26 | 0.08 | $9.84 \times 10^{-4}$ | 3885 |
| **Teen Focus4** | 17.5 | 0.37 | 0.10 | $3.27 \times 10^{-5}$ | 3373 |

$h^2$: SNP-heritability; SE: standard error; N: sample size.

**Supplementary Table 11: Estimated genetic correlations between multiple ages from cross-sectional genetic analyses in GCTA**

| Follow-up codes | chdb2 (0.8) | chdb3 (1.7) | F7 (7.6) | F10 (10.7) | T2 (13.9) | T3 (15.5) | T4 (17.5) |
|---|---|---|---|---|---|---|---|
| **chdb2** | 1 (0) | | | | | | |
| **chdb3** | 0.98 (0.11) | 1 (0) | | | | | |
| **F7** | 0.58 (0.15) | 0.68 (0.15) | 1 (0) | | | | |
| **F10** | 0.37 (0.25) | 0.31 (0.25) | 1 (0.03) | 1 (0) | | | |
| **T2** | 0.36 (0.50) | 0.66 (0.32) | 0.9 (0.07) | 0.85 (0.07) | 1 (0) | | |
| **T3** | 0.25 (0.24) | 0.55 (0.20) | 0.97 (0.07) | 0.96 (0.08) | 0.98 (0.03) | 1 (0) | |
| **T4** | 0.39 (0.20) | 0.33 (0.25) | 0.91 (0.1) | 0.94 (0.07) | 0.89 (0.06) | 1 (0.06) | 1 (0) |

The genetic correlations for corresponding cross-sectional follow-ups (as indicated by the columns and rows) are presented in the lower triangle of the table, with standard errors enclosed in brackets. Follow-up codes are: chdb2: Child health database; chdb3: Child health database 3; F7: Focus@7; F10: Focus@10; T2: Teen Focus2; T3: Teen Focus3; T4: Teen Focus4. The average age (in years) in each code is shown in brackets in the header.

**Supplementary Table 12: Estimated fixed effects from the random regression model with heterogenous errors.**

| Model Term | Level | Fixed effects | SE |
|---|---|---|---|
| Intercept | - | 2.9020 | 0.0019 |
| Legendre polynomials | slope | 0.1624 | 0.0014 |
| | $slope^2$ | 0.0531 | 0.0009 |
| | $slope^3$ | -0.0343 | 0.0007 |
| Source(clinic/questionnaire) | questionnaire | -0.0159 | 0.0007 |
| Sex(male/female) | female | -0.0111 | 0.0028 |
| Interaction terms between sex and Legendre polynomials | female*slope | -0.0203 | 0.0020 |
| | $female*slope^2$ | 0.0090 | 0.0014 |
| | $female*slope^3$ | 0.0033 | 0.0010 |

Source: measurement sources SE: standard errors of estimated fixed effect.

**Supplementary Table 13: Estimated variances and covariances for random effect terms in the random regression model heterogeneous errors.**

| Unique Individual Variance-Covariance Matrix($K_i$) | | | | |
|---|---|---|---|---|
| | intercept | slope | slope$^2$ | slope$^3$ |
| intercept | 0.0168 (0.0013) | | | |
| slope | 0.0053 (0.0005) | 0.0036 (0.0003) | | |
| slope$^2$ | -0.0030 (0.0003) | -0.0007 (0.0001) | 0.0018 (0.0001) | |
| slope$^3$ | -0.0019 (0.0001) | -0.0010 (0.0000) | 0.0006 (0.0000) | 0.0008 (0.0000) |
| **Additive Genetic Variance-Covariance Matrix($K_g$)** | | | | |
| | intercept | slope | slope$^2$ | |
| intercept | 0.0075 (0.0013) | | | |
| slope | 0.0024 (0.0005) | 0.0017 (0.0003) | | |
| slope$^2$ | -0.0012 (0.0003) | -0.0004 (0.0001) | 0.0004 (0.0001) | |
| **Residual Variances** | | | | |
| Residual 1 | 0.0017 (0.0001) | | | |
| Residual 2 | 0.0010 (0.0000) | | | |
| Residual 3 | 0.0023 (0.0001) | | | |
| Residual 4 | 0.0016 (0.0001) | | | |
| Residual 5 | 0.0038 (0.0001) | | | |

| | |
|---|---|
| Residual 6 | 0.0052 (0.0003) |
| Residual 7 | 0.0016 (0.0000) |
| Residual 8 | 0.0012 (0.0000) |
| Residual 9 | 0.0015 (0.0000) |
| Residual 10 | 0.0013 (0.0000) |
| Residual 11 | 0.0014 (0.0000) |
| Residual 12 | 0.0016 (0.0000) |
| Residual 13 | 0.0030 (0.0001) |
| Residual 14 | 0.0026 (0.0002) |
| Residual 15 | 0.0029 (0.0001) |
| Residual 16 | 0.00319 (0.0002) |
| Residual 17 | 0.00242 (0.0001) |

The variances (SE) are on the diagonals and the values below the diagonal are the covariances (SE). The residual terms represent the estimated residuals of each yearly age bins ranging from 1 to 18 years, the number indicates the starting year of the corresponding age bin.

**Supplementary Table 14: Estimated heritability of BMI at yearly intervals from one to 18 years of age from the random regression model with heterogeneous errors**

| Age (years) | $V_g$ | $Var(V_g)$ | $V_i$ | $Var(V_i)$ | $V_P$ | $Var(V_P)$ | $\sigma_e^2$ | $Var(\sigma_e^2)$ | $h^2_{SNP}$ | $SE(h^2_{SNP})$ |
|---|---|---|---|---|---|---|---|---|---|---|
| **1** | $1.85 \times 10^{-3}$ | $2.20 \times 10^{-7}$ | $4.58 \times 10^{-3}$ | $2.49 \times 10^{-7}$ | $8.16 \times 10^{-3}$ | $5.96 \times 10^{-8}$ | $1.73 \times 10^{-3}$ | $4.25 \times 10^{-9}$ | 0.227 | 0.057 |
| **2** | $1.51 \times 10^{-3}$ | $1.25 \times 10^{-7}$ | $3.33 \times 10^{-3}$ | $1.27 \times 10^{-7}$ | $5.86 \times 10^{-3}$ | $1.86 \times 10^{-8}$ | $1.02 \times 10^{-3}$ | $2.11 \times 10^{-9}$ | 0.258 | 0.060 |
| **3** | $1.54 \times 10^{-3}$ | $1.06 \times 10^{-7}$ | $3.30 \times 10^{-3}$ | $1.05 \times 10^{-7}$ | $7.14 \times 10^{-3}$ | $1.58 \times 10^{-8}$ | $2.29 \times 10^{-3}$ | $3.69 \times 10^{-9}$ | 0.216 | 0.045 |
| **4** | $1.86 \times 10^{-3}$ | $1.30 \times 10^{-7}$ | $3.86 \times 10^{-3}$ | $1.28 \times 10^{-7}$ | $7.30 \times 10^{-3}$ | $2.39 \times 10^{-8}$ | $1.58 \times 10^{-3}$ | $8.26 \times 10^{-9}$ | 0.255 | 0.049 |
| **5** | $2.37 \times 10^{-3}$ | $1.92 \times 10^{-7}$ | $4.75 \times 10^{-3}$ | $1.87 \times 10^{-7}$ | $1.09 \times 10^{-2}$ | $3.37 \times 10^{-8}$ | $3.77 \times 10^{-3}$ | $1.11 \times 10^{-8}$ | 0.217 | 0.040 |
| **6** | $3.00 \times 10^{-3}$ | $2.91 \times 10^{-7}$ | $5.95 \times 10^{-3}$ | $2.82 \times 10^{-7}$ | $1.41 \times 10^{-2}$ | $1.01 \times 10^{-7}$ | $5.20 \times 10^{-3}$ | $6.86 \times 10^{-8}$ | 0.212 | 0.038 |
| **7** | $3.68 \times 10^{-3}$ | $4.21 \times 10^{-7}$ | $7.49 \times 10^{-3}$ | $4.09 \times 10^{-7}$ | $1.28 \times 10^{-2}$ | $4.98 \times 10^{-8}$ | $1.62 \times 10^{-3}$ | $1.90 \times 10^{-9}$ | 0.288 | 0.050 |
| **8** | $4.38 \times 10^{-3}$ | $5.73 \times 10^{-7}$ | $9.41 \times 10^{-3}$ | $5.60 \times 10^{-7}$ | $1.49 \times 10^{-2}$ | $6.99 \times 10^{-8}$ | $1.15 \times 10^{-3}$ | $1.08 \times 10^{-9}$ | 0.293 | 0.050 |
| **9** | $5.03 \times 10^{-3}$ | $7.31 \times 10^{-7}$ | $1.17 \times 10^{-2}$ | $7.20 \times 10^{-7}$ | $1.82 \times 10^{-2}$ | $9.72 \times 10^{-8}$ | $1.51 \times 10^{-3}$ | $1.36 \times 10^{-9}$ | 0.276 | 0.046 |
| **10** | $5.61 \times 10^{-3}$ | $8.79 \times 10^{-7}$ | $1.41 \times 10^{-2}$ | $8.78 \times 10^{-7}$ | $2.10 \times 10^{-2}$ | $1.29 \times 10^{-7}$ | $1.28 \times 10^{-3}$ | $8.94 \times 10^{-10}$ | 0.268 | 0.044 |
| **11** | $6.10 \times 10^{-3}$ | $1.01 \times 10^{-6}$ | $1.64 \times 10^{-2}$ | $1.02 \times 10^{-6}$ | $2.39 \times 10^{-2}$ | $1.64 \times 10^{-7}$ | $1.42 \times 10^{-3}$ | $1.41 \times 10^{-9}$ | 0.255 | 0.041 |
| **12** | $6.49 \times 10^{-3}$ | $1.12 \times 10^{-6}$ | $1.82 \times 10^{-2}$ | $1.15 \times 10^{-6}$ | $2.63 \times 10^{-2}$ | $1.99 \times 10^{-7}$ | $1.57 \times 10^{-3}$ | $1.96 \times 10^{-9}$ | 0.247 | 0.040 |
| **13** | $6.78 \times 10^{-3}$ | $1.21 \times 10^{-6}$ | $1.93 \times 10^{-2}$ | $1.25 \times 10^{-6}$ | $2.91 \times 10^{-2}$ | $2.27 \times 10^{-7}$ | $3.00 \times 10^{-3}$ | $3.71 \times 10^{-9}$ | 0.233 | 0.037 |
| **14** | $6.98 \times 10^{-3}$ | $1.31 \times 10^{-6}$ | $1.92 \times 10^{-2}$ | $1.35 \times 10^{-6}$ | $2.88 \times 10^{-2}$ | $2.61 \times 10^{-7}$ | $2.61 \times 10^{-3}$ | $2.43 \times 10^{-8}$ | 0.242 | 0.039 |
| **15** | $7.11 \times 10^{-3}$ | $1.44 \times 10^{-6}$ | $1.81 \times 10^{-2}$ | $1.47 \times 10^{-6}$ | $2.81 \times 10^{-2}$ | $2.40 \times 10^{-7}$ | $2.91 \times 10^{-3}$ | $7.90 \times 10^{-9}$ | 0.253 | 0.042 |
| **16** | $7.20 \times 10^{-3}$ | $1.66 \times 10^{-6}$ | $1.61 \times 10^{-2}$ | $1.64 \times 10^{-6}$ | $2.65 \times 10^{-2}$ | $2.41 \times 10^{-7}$ | $3.19 \times 10^{-3}$ | $2.60 \times 10^{-8}$ | 0.272 | 0.048 |
| **17** | $7.30 \times 10^{-3}$ | $2.02 \times 10^{-6}$ | $1.42 \times 10^{-2}$ | $1.98 \times 10^{-6}$ | $2.39 \times 10^{-2}$ | $2.38 \times 10^{-7}$ | $2.42 \times 10^{-3}$ | $1.58 \times 10^{-8}$ | 0.305 | 0.058 |
| **18** | $7.46 \times 10^{-3}$ | $2.63 \times 10^{-6}$ | $1.42 \times 10^{-2}$ | $2.67 \times 10^{-6}$ | $2.41 \times 10^{-2}$ | $3.68 \times 10^{-7}$ | $2.42 \times 10^{-3}$ | $1.58 \times 10^{-8}$ | 0.310 | 0.067 |

$V_g$: estimated additive genetic variance; $Var(V_g)$: estimated variance of the additive genetic variance ($V_g$); $V_i$: estimated unique individual variance. $Var(V_i)$: estimated variance of unique individual variance; $V_P$: estimated phenotypic variance; $Var(V_P)$: estimated variance of phenotypic variance; $\sigma_e^2$: estimated residual variance; $Var(\sigma_e^2)$: estimated variance of residual variance. $h^2_{snp}$: estimated SNP-heritability; $SE(h^2_{snp})$: standard error of SNP-heritability estimate.

**Supplementary Notes**

**Supplementary Note 1: ASReml .as file for main analysis**

```
!MP 16 !NO GRAPHICS !WORKSPACE 32 !RENAME

ALSPAC BMI repeated measures one year to 18 years

ID 6291   !A
age
sex 2 !A
weight
height
bmi
logbmi

grm_unrel_grm_ID_noheader_1_18y_3times.grm                                   !HINV
grm_unrel_grm_ID_noheader_1_18y_3times.txt
bmigrowth_ID_1_18y_3times.dat !skip 1 !MAXIT 150

log(bmi) ~ mu sex*leg(age,-3) !r leg(age,3).ide(ID) leg(age,2).giv(ID,1)

1 1 2                    #variance header line, no. R R G
0 0 IDV 0 !S2==1
leg(age,3).ide(ID) 2
leg(age,3) 0 US 0 !GP !+10
0.2928E-01
0.8908E-02  0.6640E-02
-0.5594E-02 -0.3050E-03  0.3351E-02
-0.7722E-05 -0.7399E-03  0.2433E-03  0.1227E-02
ide(ID)

leg(age,2).giv(ID,1) 2
leg(age,2) 0 US !GP !+6
0.002
0 0.001
0 0  0.001
ID 0 GIV1
```

**Supplementary Note 2: R script for calculating 95% confidence intervals for the eigenvalues using numerical simulation**

```
#loading estimated variances from .asr file
tmp1 = read.table("ALSPAC_RRM_g2_src_v3.asr",skip=94,nrow=17)

# residual
e = tmp1[1,5]

# unique individual effects
Ki = matrix(NA, nrow=4, ncol=4)
for (i in 1:10) {
  Ki[tmp1[(i+1),3],tmp1[(i+1),4]] = tmp1[(i+1),5]
  Ki[tmp1[(i+1),4],tmp1[(i+1),3]] = tmp1[(i+1),5]
}

# genetic effects
Kg = matrix(NA, nrow=3, ncol=3)
for (i in 1:6) {
  Kg[tmp1[(i+11),3],tmp1[(i+11),4]] = tmp1[(i+11),5]
  Kg[tmp1[(i+11),4],tmp1[(i+11),3]] = tmp1[(i+11),5]
}

# vvp estimated variance of variance components
# tmp2 = scan("ALSPAC_RRM_g2_v3.vvp",skip=1)
tmp2 = scan("ALSPAC_RRM_g2_src_v3.vvp",skip=1)
vvp = diag(0,nrow=17)
vvp[lower.tri(vvp,diag=T)] = tmp2
vvp[upper.tri(vvp,diag=T)] = tmp2
V = vvp[12:17,12:17] #estimated variance of the additive genetic variance component

#########################
# eigenvalue decomposition of Kg
#########################
Kg
# [,1]        [,2]        [,3]
# [1,]  0.00734918  0.002429220 -0.001162300
# [2,]  0.00242922  0.001728870 -0.000392931
# [3,] -0.00116230 -0.000392931  0.000400509
PCs = eigen(Kg)

# confidence limits for eigenvalues
library(ggplot2)
gHat = Kg[lower.tri(Kg,diag=T)]
# Cholesky decomposition
L = chol(V)
nvars = dim(V)[1]
```

```
simGcov = matrix(NA,nrow=6000,ncol=6)
simValues = matrix(NA,nrow=6000,ncol=3)
for (i in 1:6000) {
  simGcov[i,] = gHat + t(L) %*% matrix(rnorm(nvars))
  simG = matrix(NA,nrow=3, ncol=3)
  simG[lower.tri(simG,diag=T)] = simGcov[i,]
  simG[upper.tri(simG)] = simG[lower.tri(simG)]
  simValues[i,] = eigen(simG)$values
}

data1 = data.frame(values = c(simGcov), component = sort(rep(1:6,6000)))
data2 = data.frame(values = c(simValues), eigenValue = sort(rep(1:3,6000)))
est1 = data.frame(values = gHat, component = 1:6)
est2 = data.frame(values = PCs$values, eigenValue = 1:3)

# plot estimates with simulated ci
estimate = PCs$values
l.ci =  apply(simValues,2,quantile,0.025)
u.ci =  apply(simValues,2,quantile,0.975)

data3 = data.frame(eigenValue = paste0("eigenValue",1:3),estimate,l.ci,u.ci)
ggplot(data3) + geom_bar(aes(x=eigenValue, y=estimate), stat="identity", fill="skyblue") +
  geom_errorbar(aes(x=eigenValue, ymin=l.ci, ymax=u.ci), alpha=0.9, width = 0.1, size = 
1.1, col="orange") +
  xlab("") + ylab("")
```

**Supplementary Note 3: ASReml .as file for adjusting for adult BMI PGS**

```
!MP 16 !NO GRAPHICS !WORKSPACE 32 !RENAME

ALSPAC BMI repeated measures one year plus

ID 6291   !A
age
sex 2 !A
weight
height
bmi
source 2 !A
SCORESUM #PGS
logbmi

grm_unrel_grm_ID_noheader_1_18y_3times_v4.grm !HINV
grm_unrel_grm_ID_noheader_1_18y_3times_v4.txt
bmigrowth_ID_1_18y_3times_v4.dat !skip 1 !MAXIT 150 !ASUV

log(bmi) ~ mu sex*leg(age,-3) source SCORESUM*leg(age,-3) !r leg(age,3).ide(ID)
leg(age,2).giv(ID,1)

1 1 2                #variance header line, no. R R G
0 0 IDV 0 !S2==1
leg(age,3).ide(ID) 2
leg(age,3) 0 US 0 !GP !+10
  0.2278E-01
  0.7486E-02  0.5335E-02
 -0.3828E-09 -0.8910E-09  0.2167E-02
 -0.4635E-09 -0.5613E-03  0.6141E-03  0.7907E-03
ide(ID)

leg(age,2).giv(ID,1) 2
leg(age,2) 0 US !GP !+6
0
0 0
0 0 0
ID 0 GIV1
```

**Supplementary Note 4: ASReml .as file for heterogeneity analysis**

```
!MP 16 !NO GRAPHICS !WORKSPACE 32 !RENAME

ALSPAC BMI repeated measures one year plus

ID 6291   !A
age
sex 2 !A
weight
height
bmi
source 2 !A
logbmi

grm_unrel_grm_ID_noheader_1_18y_3times_v3.grm                                   !HINV
grm_unrel_grm_ID_noheader_1_18y_3times_v3.txt
bmigrowth_ID_1_18y_3times_v3.dat !skip 1 !MAXIT 150 !ASUV

log(bmi) ~ mu sex*leg(age,-3) source !r leg(age,3).ide(ID) leg(age,2).giv(ID,1) !f mv

17 1 2  # !STEP .01
5175 0  ID # !S2==1
2189 0  ID # !S2==1
5223 0  ID # !S2==1
1166 0  ID # !S2==1
3479 0  ID # !S2==1
969 0  ID # !S2==1
5190 0  ID # !S2==1
4563 0  ID # !S2==1
5040 0  ID # !S2==1
6241 0  ID # !S2==1
4903 0  ID # !S2==1
4503 0  ID # !S2==1
7069 0  ID # !S2==1
914 0  ID # !S2==1
3618 0  ID # !S2==1
1893 0  ID # !S2==1
3794 0  ID # !S2==1

leg(age,3).ide(ID) 2
leg(age,3) 0 US 0 !GP !+10
0.1650E-01
0.5330E-02  0.3516E-02
-0.2808E-02 -0.5955E-03  0.1684E-02
-0.1685E-02 -0.9866E-03  0.5489E-03  0.7707E-03
ide(ID)
```

```
leg(age,2).giv(ID,1) 2
leg(age,2) 0 US !GP !+6
0.7348E-02
0.2429E-02  0.1729E-02
-0.1162E-02 -0.3929E-03  0.4005E-03
ID 0 GIV1
```

**Supplementary Note 5: Eigenvalue decomposition of genetic correlation matrix of BMI from the previously published COADTwins project**

We performed eigenvalue decomposition using the genetic correlation matrix of BMI obtained from the previously published COADTwins project [1]. By doing this, we aimed to validate the pattern of genetic inheritance by estimating $\mathbf{K_g}$ using the reported genetic correlation matrix and heritability across different ages from the CODATwins project. This should give similar results to the decomposition of $\mathbf{K_g}$ given that the age ranges included in Silventoinen is similar to the current study. Silventoinen *et al.* (2022) used Cholesky decomposition (i.e. multivariate ACE model, i.e. a common statistical model in twin and adoption studies, which breaks down phenotypic variance into three parts: additive genetic [A], shared environment [C], and specific environment plus measurement error [E]) on twin cohorts and reported correlation matrices of additive genetic components for both genders from 1 to 18 years [1]. We converted the correlation matrix to a covariance matrix (equivalent to $\mathbf{V_g}$; an 18 by 18 matrix) using the estimated standard deviations of the additive genetic component at each age. Since they did not provide estimates of heritability ($h^2$) or additive genetic variance, we approximated the genetic variance by using the phenotypic variance of BMI for each age and the previously reported heritability from CODATwins [2].

$$\widehat{v_g} = \widehat{\boldsymbol{v_p}}\widehat{h^2} = \widehat{sd_p^2}\widehat{h^2}$$
$$\widehat{sd_g} = \sqrt{\widehat{v_g}} \qquad \text{Equation 1}$$
$$\widehat{cov_g(\mathrm{t_1},\mathrm{t_2})} = \widehat{r_g(\mathrm{t_1},\mathrm{t_2})}\ \widehat{sd_{g1}}\ \widehat{sd_{g2}}$$

Where $\widehat{v_g}$ is the estimated genetic variance of a given age, $\widehat{v_p}$ is the estimated phenotypic variance, $\widehat{h^2}$ is estimated heritability in twin study, $\widehat{sd_g}$ and $\widehat{sd_p}$ are estimated standard deviations of genetic and phenotypic variances, respectively, $\widehat{cov_g(\mathrm{t_1},\mathrm{t_2})}$ and $\widehat{r_g(\mathrm{t_1},\mathrm{t_2})}$ are estimated genetic covariance and correlation between ages $t_1$ and $t_2$, respectively. $\widehat{sd_{g1}}$ and $\widehat{sd_{g2}}$ are estimated standard deviations of genetic variances at time points $t_1$ and $t_2$, respectively. This covariance matrix was transformed into the covariance matrix of polynomials ($\boldsymbol{\Phi}^+$, approximation of $\mathbf{K_g}$, a 3 x 3 matrix) through the pseudo-inverse [3] of the $\boldsymbol{\Phi}$ matrix (Legendre coefficient matrix, 18 x 3 matrix, Equation 2 and 6[4]). The pseudo inverse matrix, $\boldsymbol{\Phi}^+$, can be calculated as follows.

$$\boldsymbol{\Phi}^+ = (\boldsymbol{\Phi}'\boldsymbol{\Phi})^{-1}\boldsymbol{\Phi}'$$
$$\hat{\mathbf{V}}_{\mathbf{g}} = \boldsymbol{\Phi}\boldsymbol{\Phi}^+\boldsymbol{\Phi}' \qquad \text{Equation 2}$$

where $\hat{\mathbf{V}}_{\mathbf{g}}$ is of order $t \times t$, where t are ages across the BMI trajectory of interest for evaluation.

Finally, we used the eigen() function in R to perform the eigenvalue decomposition on the polynomial covariance matrix and visualised the eigenfunctions in comparison to ours (**Supplementary Figure 9**).

References

1. Silventoinen K, Li W, Jelenkovic A, et al. Changing genetic architecture of body mass index from infancy to early adulthood: an individual based pooled analysis of 25 twin cohorts. *Int J Obes (Lond)* 2022; **46**: 1901-9.
2. Silventoinen K, Jelenkovic A, Sund R, et al. Genetic and environmental effects on body mass index from infancy to the onset of adulthood: an individual-based pooled analysis of 45 twin cohorts participating in the COllaborative project of Development of Anthropometrical measures in Twins (CODATwins) study. *Am J Clin Nutr* 2016; **104**: 371-9.
3. Penrose R. A generalized inverse for matrices. *Mathematical Proceedings of the Cambridge Philosophical Society* 1955; **51**: 406-13.
4. Kirkpatrick M, Lofsvold D, Bulmer M. Analysis of the inheritance, selection and evolution of growth trajectories. *Genetics* 1990; **124**: 979-93.
